# Socioeconomic deprivation and risk of early-onset pre-eclampsia in England: a national population-based cohort study

**DOI:** 10.64898/2026.07.01.26355976

**Authors:** Ethan Phillips, Federica Caretta Cortegiani, Catherine Aiken, Marian Knight, Harshita Kajaria-Montag, Agni Orfanoudaki, Yueyang Zhong

## Abstract

**Objectives:** To examine the association between small-area socioeconomic deprivation and risk of early-onset pre-eclampsia (diagnosed <34 weeks gestation) in England, and to assess the relative contributions of individual-level risk factors and variation between maternity care sites to observed inequalities.

**Design:** Retrospective population-based cohort study.

**Setting:** National Health Service (NHS)-funded maternity services in England between 1 January 2021 and 31 March 2025.

**Participants:** 1,027,707 nulliparous pregnant women aged 13-60 years receiving NHS-funded maternity care in England with singleton pregnancies and non-missing deprivation data. Secondary analyses were conducted for 940,505 multiparous pregnant women.

**Main outcome measures:** Early-onset pre-eclampsia, defined as diagnosis before 34 completed weeks of gestation.

**Results:** Increasing socioeconomic deprivation was associated with higher odds of early-onset pre-eclampsia among nulliparous women across all regression models. In the confounder-adjusted model, each one-point increase in the continuous deprivation score (scaled 0–10) was associated with a 3.4% increase in odds of early-onset pre-eclampsia (adjusted odds ratio (aOR) 1.034, 95% confidence interval (CI) 1.027 to 1.041). Adjustment for theorized mediators attenuated the association modestly (aOR 1.023, 95% CI 1.017 to 1.030), while additional adjustment for hospital site further attenuated the association (aOR 1.016, 95% CI 1.009 to 1.023). Elevated BMI, circulatory disease, maternal age over 40 years, Black ethnicity, and endocrine/metabolic disease were among the strongest predictors of early-onset pre-eclampsia. Similar but stronger deprivation associations were observed among multiparous women. Associations between deprivation and late-onset pre-eclampsia were comparatively weak or absent after adjustment.

**Conclusions:** Socioeconomic deprivation was associated with increased risk of early-onset pre-eclampsia in England, particularly among multiparous women. Both individual-level risk factors and variation between maternity care sites appeared to contribute to observed inequalities. These findings support the importance of combining targeted clinical risk reduction with efforts to reduce unwarranted variation in NHS maternity care delivery.

**What is already known on this topic**
Socioeconomic deprivation is associated with significantly worse maternal outcomes. Pre-eclampsia affects 2-8% of pregnant women in England, with early-onset and late-onset subtypes thought to have partly distinct aetiologies. The association between socioeconomic deprivation and pre-eclampsia has not been examined separately by onset subtype or maternal parity.
**What this study adds**
Socioeconomic deprivation was associated primarily with early-onset rather than late-onset pre-eclampsia. The association persisted after adjustment for demographic and clinical risk factors, but attenuated following adjustment for hospital site, suggesting that both individual-level and health-system factors contribute to observed inequalities. Use of a continuous neighborhood deprivation measure demonstrated a graded deprivation-risk relationship that was robust across multiple model specifications. These findings support the need for both targeted clinical interventions among deprived populations and structural efforts to standardize NHS maternity care quality across sites.

## Introduction

In England, the National Health Service (NHS) offers maternity services to all pregnant women, regardless of citizenship or immigration status [1], thereby seeking to prevent maternal and early-childhood health inequities on the basis of income. Even so, significant inequalities in maternal outcomes persist [2]. Socioeconomic deprivation, encompassing multi-dimensional disadvantage across income, employment, education, health exposure and disability, crime, housing, and lived environment [3], has consistently been shown to strongly correlate with worse maternal and neonatal health outcomes [4, 5]. From 2022 to 2024, women living in the most deprived areas of the United Kingdom (UK) experienced maternal mortality at almost twice the rate of women in the most well-off areas [5]. Pregnant women of non-White ethnicity, particularly Black women, also experience significantly worse health outcomes compared to their White peers in England [5]. In 2024, nearly half of NHS maternity sites were rated “inadequate” or “requires improvement” by the Care Quality Commission (CQC), highlighting stark variation in care quality across locations [6]. These disparities have recently drawn national attention, leading to an independent investigation into NHS maternity services [7]. Socioeconomic deprivation may influence maternal outcomes through both individual-level pathways, such as lifestyle, comorbidities, and health behaviors, and through healthcare system factors, including the quality and accessibility of services received [8]. While individual-level factors may be shaped by means and opportunity, they can be ameliorated through public health and clinical intervention. Healthcare system factors, however, require more complex organizational and system interventions or policy change. Identifying the primary loci and pathways through which deprivation influences maternal and neonatal health is critical, as individual-level and system-level determinants call for fundamentally different responses.

Pre-eclampsia is one condition through which deprivation may be particularly consequential, given its sensitivity to both individual-level risk factors and the quality of antenatal care received, thus making it an informative lens through which to examine health inequities in maternity care. Pre-eclampsia is a serious pregnancy-related hypertensive disorder that is responsible for significant maternal morbidity and mortality [9, 10]. While the condition commonly presents as hypertension in combination with proteinuria, disease progression can lead to systemic inflammation, hemorrhage or other blood disorders, and organ dysfunction/failure (especially involving the liver, kidneys, heart, or brain) [9, 10]. In England, pre-eclampsia is diagnosed in approximately 2–8% of pregnancies, with prevalence being higher among women in their first pregnancy compared to subsequent pregnancies [11, 12]. Despite robust clinical evidence on the impacts and treatment of pre-eclampsia, its causes are poorly understood [9, 10]. The condition is commonly subdivided by gestational age at presentation, with diagnoses before 34 weeks classified as early-onset and those at or after 34 weeks classified as late-onset [13, 14]. The two subtypes are thought to have partly distinct pathophysiological mechanisms, with early-onset disease more strongly linked to defective placentation and impaired uteroplacental perfusion, and late-onset disease more closely associated with maternal cardiovascular and metabolic predisposition [13, 14]. Early-onset pre-eclampsia is associated with substantially greater maternal and neonatal morbidity and frequently requires intensified antenatal monitoring, preterm delivery, and neonatal intensive care support [13–15]. Despite these distinctions, most epidemiological studies continue to analyze pre-eclampsia as a single outcome, potentially obscuring important heterogeneity in risk profiles and underlying mechanisms.

The relationship between socioeconomic deprivation and pre-eclampsia has not been well characterized separately by onset subtype or maternal parity. This distinction may be particularly important among nulliparous women, for whom prior obstetric history is unavailable and risk stratification therefore relies more heavily on demographic, clinical, and social characteristics. By contrast, prior pre-eclampsia is already known to be one of the strongest predictors of recurrence among multiparous women [20]. This study therefore aimed primarily at examining the association between neighborhood socioeconomic deprivation and early-onset pre-eclampsia among nulliparous women in England. Secondary aims were to assess the relative contributions of theorized individual-level mediators and variation between maternity care sites to observed inequalities, and to compare these findings with corresponding analyses among multiparous women and for late-onset disease.

## Methods

### Data sources

We conducted a retrospective population-based cohort study of pregnancies recorded in NHS-funded maternity services in England between 1 January 2021 and 31 March 2025. The primary data source was the NHS Maternity Services Data Set (MSDS) version 2.0, a national administrative dataset capturing patient-level information at key stages of the maternity care pathway [16]. MSDS data are submitted to NHS England by all NHS-funded maternity units from the first antenatal booking appointment (approximately 8 weeks after the last menstrual period) through service discharge in the weeks following delivery [16]. Not all units submitted complete data during the study period, resulting in fewer MSDS records than live births reported nationally by the Office for National Statistics.

MSDS records were linked to the Emergency Care Data Set (ECDS) [17] and to Hospital Episode Statistics (HES) [18] using each woman’s pseudonymized NHS number. ECDS provided coded diagnoses from attendances at NHS accident and emergency (A&E) departments, used to ascertain maternal comorbidities before and during pregnancy. HES provided inpatient admission records, used both to supplement missing values in the MSDS and to identify postpartum readmissions.

Anonymized MSDS, ECDS, and HES data were accessed through a secure data environment under a data sharing agreement with NHS England. Reporting follows the Strengthening the Reporting of Observational Studies in Epidemiology (STROBE) guidelines.

### Study population

We included all pregnant women recorded in the MSDS with an estimated date of delivery between 1 January 2021 and 31 March 2025; MSDS records starting from 1 April 2018 were retained only to ascertain prior obstetric history and covariates, but were not included in the regression analysis due to higher data missingness. Notably, MSDS v2.0 coverage of NHS-funded deliveries did not reach 94% until 2021/22 [19]. We excluded pregnancies from mothers aged under 13 or over 60 at booking, multifetal births or missing number of births, those with missing IMD decile or inconsistent linkage between birth date and admission to labor and delivery, and, where a woman had multiple records, all but her earliest recorded pregnancy in the dataset.

The cohort was then stratified into two groups based on parity: nulliparous women (no prior pregnancy lasting >24 weeks) and multiparous women (at least one prior pregnancy lasting >24 weeks). Parity was determined on the basis of the full obstetric history, so as to accurately categorize each woman’s parity even where the prior pregnancy was not included within the temporal scope of the cohort. This split was pre-specified given the strong modifying effect of parity on pre-eclampsia risk. Within the nulliparous group, pregnancies with a recorded prior diagnosis of pre-eclampsia or gestational diabetes mellitus were additionally excluded, as such records are clinically inconsistent with nulliparity and likely reflect data quality issues. We chose to focus primarily on nulliparous women given their higher prevalence of pre-eclampsia and greater uncertainty of clinical risk compared to multiparous women due to lack of prior obstetric history. In multiparous women, history of pre-eclampsia (or lack thereof) is highly informative of risk in future pregnancies [20]. Socioeconomic and demographic factors are therefore most relevant for informing clinical risk stratification among nulliparous women. A more detailed explanation of the exclusion process, including a diagram of the full data pipeline (Supplementary Figure 1), can be found in Appendix A. Results for multiparous women are also reported in the supplementary material (Appendix D).

### Exposure

The exposure of interest was area-level socioeconomic deprivation, measured using the 2019 English Indices of Multiple Deprivation (IMD). It constitutes the government’s composite index of relative deprivation for every lower-layer super output area (LSOA) in England, with each LSOA covering a neighborhood of approximately 1,500 residents [3]. Each woman was assigned an IMD score based on her residential postcode at the booking antenatal appointment. The original IMD ranking ranges from 1 (most deprived neighborhood) to 32,844 (least deprived). For interpretability, we inverted and rescaled this to a continuous deprivation score from 0.0 (least deprived) to 10.0 (most deprived), such that higher values indicate greater deprivation throughout. As a robustness check, the fully adjusted model was additionally re-estimated using a dichotomized exposure, classifying women in the most deprived 30% of neighborhoods (original IMD deciles 1–3) as deprived and all others (deciles 4–10) as non-deprived. The threshold of 30% was chosen to retain sufficient events in the exposed group for stable estimation of the early-onset pre-eclampsia association given its low population prevalence. To further assess sensitivity to this threshold choice, the model was additionally re-estimated across alternative IMD thresholds ranging from decile 1 to decile 5. Adjusted odds ratios under alternative thresholds, estimated with and without hospital-site random effects, are reported in Supplementary Figures 3 and 4 respectively (Appendix G), and remained stable across specifications.

### Outcomes

We considered early-onset pre-eclampsia the primary study outcome, distinguished from late-onset by gestational age at first qualifying diagnosis. Results for late-onset diagnoses are reported in the supplementary material (Appendix E). Pre-eclampsia during pregnancy was ascertained by combining coded diagnoses with supporting clinical findings recorded between the estimated date of conception and discharge from labor and delivery care. Coded diagnoses were identified from pregnancy-related entries in MSDS, from HES Admitted Patient Care records, and from ECDS attendances, using curated lists of ICD-10 and SNOMED CT codes provided in the Supplementary Table 1 (Appendix A). A case was further confirmed only where the coded diagnosis was accompanied by either (i) at least two recorded hypertension events or (ii) at least one hypertension and one proteinuria event during the same pregnancy, drawn from MSDS clinical findings.

Gestational age at diagnosis was calculated as the interval between the earliest qualifying diagnosis date and the estimated start of pregnancy, defined as 40 weeks before the estimated date of delivery recorded at the dating ultrasound scan. Diagnoses before 34 completed weeks of gestation were classified as early-onset; those at or after 34 weeks as late-onset [13, 14].

## Covariates

Covariates were selected according to a pre-specified directed acyclic graph (DAG) presented in Figure 1, separating variables into potential confounders and potential mediators of the deprivation–pre-eclampsia association [30]. Theorized confounders included maternal age at booking (<35 years [reference], 35–40, >40) [21, 22], self-reported ethnicity (White [reference], Black, South Asian, Mixed, or Other), and English-language proficiency recorded at booking (English speaker versus not).

**Figure 1.**
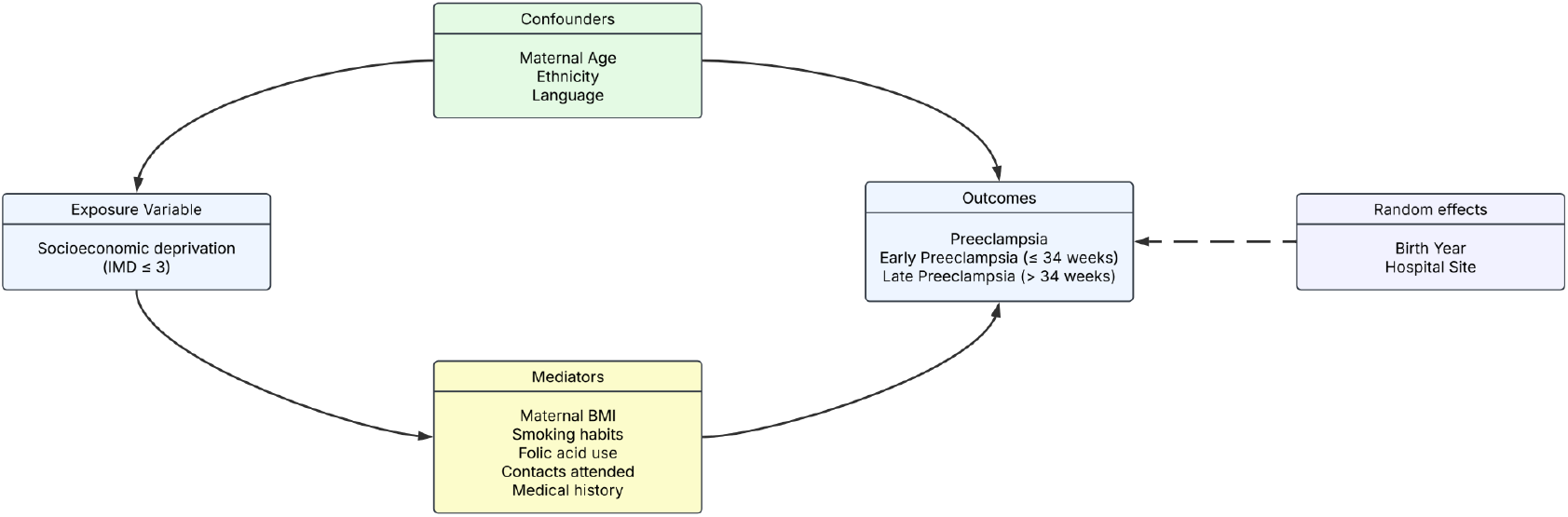
DAG showing the hypothesized causal structure linking socioeconomic deprivation to early-onset and late-onset pre-eclampsia. Arrows indicate hypothesized causal pathways.

Mediators comprised maternal body mass index (BMI); ever-smoking status; folic acid supplementation during pregnancy; the proportion of scheduled antenatal contacts attended; and binary indicators for recorded diagnoses of endocrine/metabolic, circulatory, respiratory, gastrointestinal, nervous system, musculoskeletal, and mental health conditions, as well as congenital malformations or abnormalities. BMI was calculated using maternal weight recorded at booking and an assumed average female height of 1.62 m [41], as maternal height was not consistently recorded in MSDS. Implications of this approximation are considered in the Discussion. Comorbidity diagnoses were ascertained from MSDS, HES, and ECDS records up to the end of pregnancy as described in the Appendix A (Stage 5.2). While folic acid supplementation is not consistently associated with modified risk of pre-eclampsia, it was included as a mediator to proxy for unmeasured factors such as planned pregnancy, level of patient engagement with maternal health care, and individual adherence to recommended clinical practice. Among multiparous women, prior pre-eclampsia and prior gestational diabetes diagnoses were additionally included.

### Statistical analysis

Associations between socioeconomic deprivation and each pre-eclampsia outcome (early-onset, late-onset) were estimated using hierarchical mixed-effects logistic regression, fitted separately within each parity group (nulliparous, multiparous) [31, 32]. All variables included in the regression models, together with their data type, are defined in Supplementary Table 2 (Appendix B). Three nested models were specified, adding covariates progressively in line with the DAG: model 1 contained the exposure alone; model 2 additionally adjusted for confounders (age, ethnicity, language); model 3 additionally adjusted for mediators (BMI, smoking, folic acid, antenatal contact attendance, and recorded comorbidities, plus prior pre-eclampsia and prior gestational diabetes in the multiparous group). Nested-model comparisons are interpreted as descriptive attenuation, not formal mediation.

Covariates in MSDS were, where incomplete, supplemented using linked HES records; remaining missingness was addressed using k-nearest-neighbors imputation [33–35] within strata defined by birth year and IMD decile, prior to regression fitting, as described in Appendix A (Stage 6). The missingness pattern across all included covariates before and after imputation is reported in Supplementary Table 3 (Appendix C).

Birth year was included as a random effect in all models to absorb secular variation in recorded pre-eclampsia prevalence, which we attribute primarily to changes in diagnostic coding and submission completeness over the study window rather than to true shifts in disease incidence. Each of models 1–3 was fitted in two specifications: with birth year as the sole random effect, and with birth year and hospital site as crossed random effects. Including hospital site–that is, the site of labor and delivery–as a random effect allowed us to assess whether the deprivation association was driven by between-hospital differences in care or existed independently of where women were treated. For early-onset pre-eclampsia in particular, delivery site may partly be downstream of the clinical course: women with high-risk conditions may be referred or transferred to tertiary centres, meaning that delivery site reflects not only differences in hospital quality but also referral patterns, case mix, geography, and capacity. These sources of between-hospital heterogeneity are absorbed by the hospital-level random effect, rather than attributed to individual patient characteristics.

As robustness checks, Cox proportional hazards models were fitted in parallel for both outcomes, parity groups, and random-effect specifications, treating gestational age at diagnosis as a time-to-event outcome. Kaplan–Meier survival curves were plotted for each parity group and for the full cohort, stratified by deprivation status, to visualise the gestational timing of diagnostic onset.

All analyses were conducted in Python 3.12.3 [36] within Databricks notebooks, using PySpark 4.1.0 [37, 38] for data processing and generalized linear mixed-effects logistic regression. Cox proportional hazards models and Kaplan–Meier survival curves were estimated in R 4.2.3 (RStudio) [39] using the “survival” package [40]. Confidence intervals are presented at the 95% level throughout.

### Patient and public involvement

Patients and members of the public were not directly involved in the design, conduct, or interpretation of this study. The research question was motivated by persistent evidence of socioeconomic and ethnic disparities in maternal outcomes in England, including findings from MBRRACE-UK maternal mortality surveillance [5], the Care Quality Commission’s national review of maternity services [6], and the ongoing independent investigation into NHS maternity and neonatal services [7], each of which has raised concerns shared by affected communities. We plan to disseminate findings to policy and clinical audiences through peer-reviewed publication and through engagement with NHS England, alongside other relevant maternity and women’s health networks.

## Results

### Participant cohort

The final analytical cohort comprised 1,968,212 pregnant women, including 1,027,707 nulliparous women and 940,505 multiparous women. Where a woman had more than one recorded pregnancy in the source data, only the first was retained, ensuring that each woman contributed at most one pregnancy to the cohort. Cohort derivation is shown in Figure 2.

**Figure 2.**
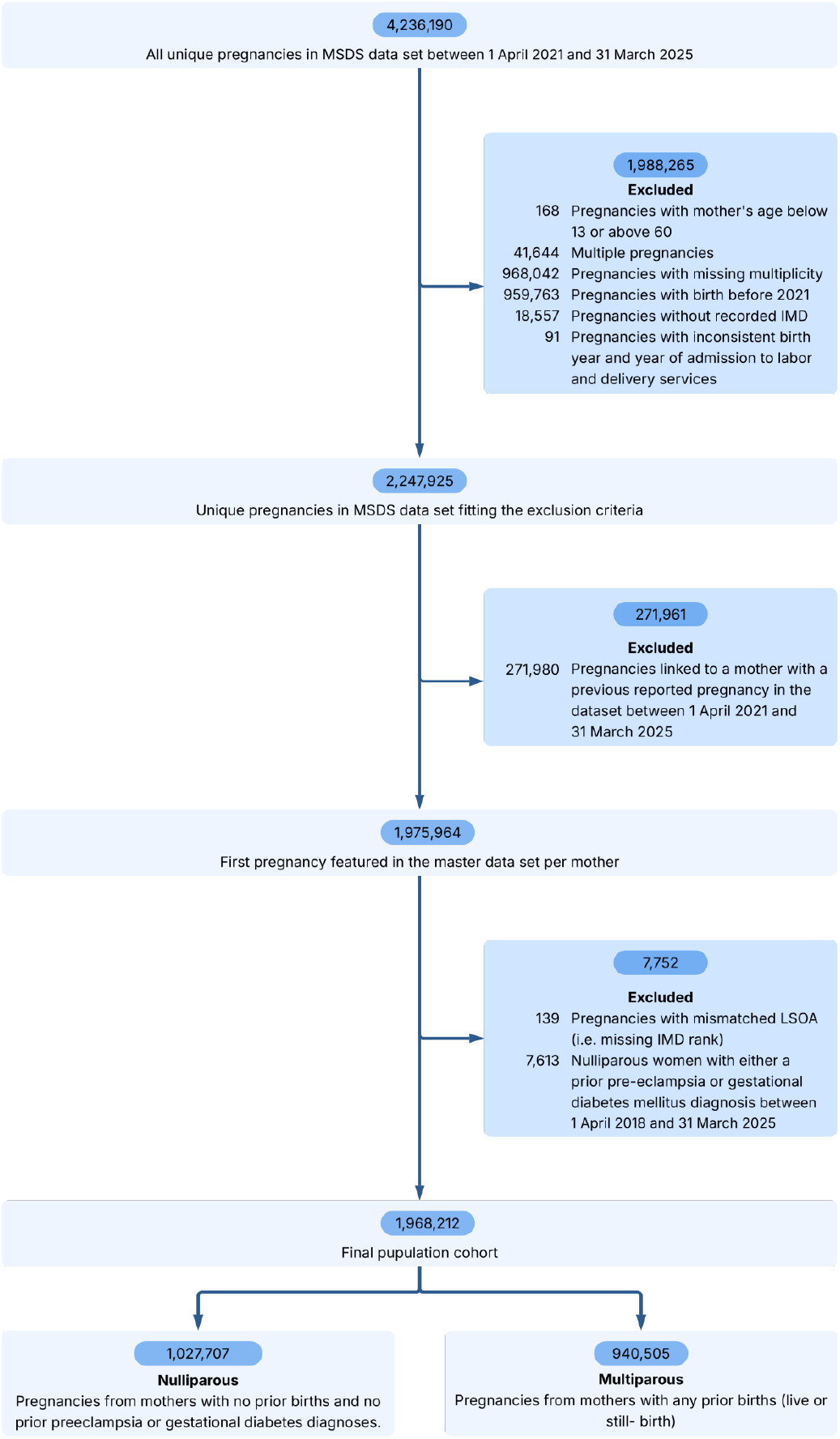
Patient exclusion diagram and parity-based cohort derivation.

Baseline characteristics of the nulliparous and multiparous cohorts are summarized in Table 1. Both groups were broadly similar in ethnic composition (White ethnicity 72% of both) and in antenatal contact attendance (81%). As expected, multiparous women were on average older (mean 31.7 versus 28.7 years). Multiparous women also had higher mean BMI and greater prevalence of obesity, smoking history, and recorded comorbidities. All categories of recorded pre-existing medical conditions were more prevalent among multiparous women, possibly due to the noted differences in mean age and BMI. These differences likely reflect both age-related accumulation of chronic disease and greater prior healthcare engagement among women with previous pregnancies, leading to more opportunities for diagnosis.

**Table 1.** Baseline characteristics of the study cohort by parity group.

| Variable | Nulliparous (n=1,027,707) | Multiparous (n=940,505) |
| --- | --- | --- |
| **Regressors** |  |  |
| Socioeconomic deprivation | 5.39 (2.81) | 5.65 (2.9) |
| Maternal age | 28.72 (5.59) | 31.69 (5.08) |
| under 35 (reference) | 876,100 (85.2%) | 657,986 (70.0%) |
| 35–40 | 125,046 (12.2%) | 229,362 (24.4%) |
| over 40 | 26,561 (2.6%) | 53,157 (5.7%) |
| Ethnicity |  |  |
| White (Reference) | 723,404 (70.4%) | 654,869 (69.6%) |
| Black | 46,966 (4.6%) | 56,671 (6.0%) |
| South Asian | 117,184 (11.4%) | 110,287 (11.7%) |
| Mixed ethnicity | 28,488 (2.8%) | 24,806 (2.6%) |
| Other ethnicity | 80,394 (7.8%) | 72,994 (7.8%) |
| Preferred Language not English | 50,252 (4.9%) | 48,332 (5.1%) |
| Maternal BMI | 27.86 (5.7) | 28.59 (5.85) |
| BMI < 18.5 | 21,307 (2.1%) | 12,736 (1.4%) |
| BMI 18.5–24.9 (reference) | 248,073 (24.1%) | 187,915 (20.0%) |
| BMI 25.0–29.9 | 202,900 (19.7%) | 185,569 (19.7%) |
| BMI > 30.0 | 202,895 (19.7%) | 220,262 (23.4%) |
| Ever Smoker | 100,604 (9.8%) | 118,270 (12.6%) |
| Folic acid during pregnancy | 884,509 (86.1%) | 782,213 (83.2%) |
| Contacts attended percentage | 81.48 (38.22) | 81.01 (38.37) |
| **Medical History** |  |  |
| Previous pre-eclampsia | — | 57,658 (6.1%) |
| Previous gestational DM | — | 49,553 (5.3%) |
| Endocrine/Metabolic Diseases | 50,721 (4.9%) | 174,094 (18.5%) |
| Mental Disorders | 122,940 (12.0%) | 220,466 (23.4%) |
| Nervous System Diseases | 35,183 (3.4%) | 51,532 (5.5%) |
| Circulatory Diseases | 35,508 (3.5%) | 53,600 (5.7%) |
| Respiratory Diseases | 96,780 (9.4%) | 143,351 (15.2%) |
| Gastrointestinal Diseases | 139,370 (13.6%) | 170,447 (18.1%) |
| Musculoskeletal Diseases | 43,611 (4.2%) | 78,217 (8.3%) |
| Malformations/Abnormalities | 7,031 (0.7%) | 7,688 (0.8%) |
| **Outcomes** |  |  |

Table 1. Baseline characteristics of the study cohort by parity group.
|  |  |  |
| --- | --- | --- |
| Pre-eclampsia | 72,663 (7.1%) | 44,613 (4.7%) |
| Early Pre-eclampsia (< 34 weeks) | 13,112 (1.3%) | 14,526 (1.5%) |
| Late Pre-eclampsia (≥ 34 weeks) | 59,437 (5.9%) | 30,042 (3.2%) |

Note: Values are prevalences (%) for binary variables and means for continuous variables (Socioeconomic deprivation, Maternal Age, Maternal BMI, Contacts attended percentage). “—” indicates the variable does not apply to that group.

Overall, among both nulliparous and multiparous women, pre-eclampsia was diagnosed in 117,276 pregnancies (6.0%), of which 27,638 (1.4%) were classified as early-onset and 89,479 (4.6%) as late-onset. 159 diagnosed cases could not be classified as early- or late-onset because the reference date was missing; these cases were excluded from the onset-specific analyses. Early-onset disease occurred at similar prevalence across parity groups (1.3% among nulliparous women versus 1.5% among multiparous), whereas late-onset disease was substantially more common among nulliparous women (5.9% versus 3.2%).

Figure 3 illustrates the relationship between neighborhood deprivation and prevalence of early-onset pre-eclampsia across the full IMD distribution. A graded increase in prevalence was observed with increasing deprivation, particularly among multiparous women. Table 2 presents regression results for early-onset pre-eclampsia among nulliparous women, the primary analysis of this study. The first three columns show models including birth year as the sole random effect, while the final column additionally includes hospital site as a random effect.

**Figure 3.**
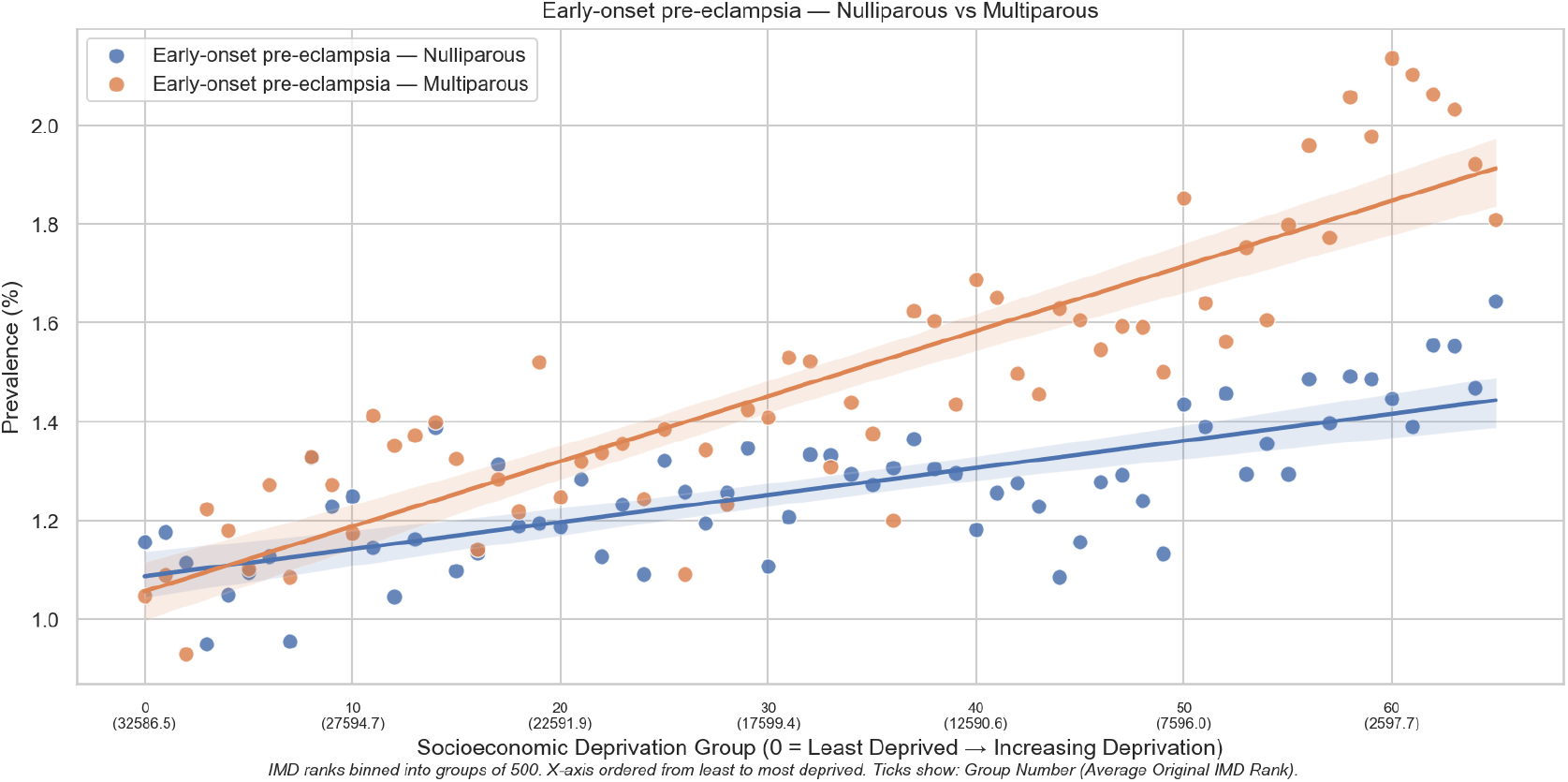
Scatterplot of the prevalence of early-onset pre-eclampsia among nulliparous and multiparous women across neighborhood socioeconomic deprivation. Deprivation is assessed at the Lower-layer Super Output Area (LSOA) level. The x-axis displays socioeconomic deprivation groups derived by inverting the national Index of Multiple Deprivation (IMD) ranks and binning them into sequential increments of 500 LSOAs. Group 0 represents the least deprived segment of the population, with higher group numbers indicating progressively greater relative deprivation. Axis ticks are formatted as n(m), where n is the sequential deprivation group number and m is the average original national IMD rank for that specific group (where 1 is the most deprived and 32,844 is the least deprived).

**Table 2.**
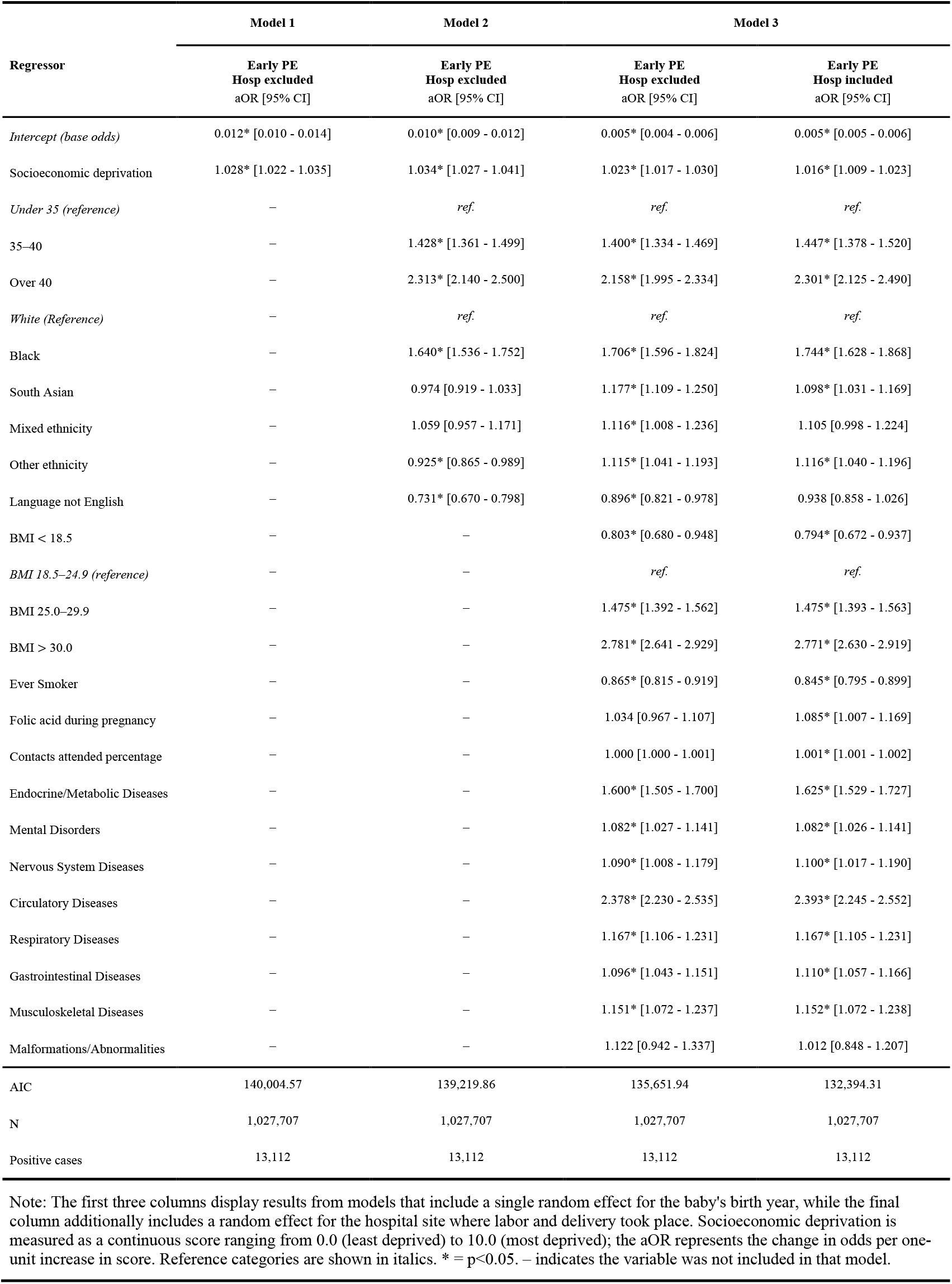
Regression results for early-onset pre-eclampsia in nulliparous women.

| Regressor | Model 1 | Model 2 | Model 3 |  |
| --- | --- | --- | --- | --- |
|  | Early PE<br>Hosp excluded<br>aOR [95% CI] | Early PE<br>Hosp excluded<br>aOR [95% CI] | Early PE<br>Hosp excluded<br>aOR [95% CI] | Early PE<br>Hosp included<br>aOR [95% CI] |
| *Intercept (base odds)* | 0.012* [0.010 - 0.014] | 0.010* [0.009 - 0.012] | 0.005* [0.004 - 0.006] | 0.005* [0.005 - 0.006] |
| Socioeconomic deprivation | 1.028* [1.022 - 1.035] | 1.034* [1.027 - 1.041] | 1.023* [1.017 - 1.030] | 1.016* [1.009 - 1.023] |
| *Under 35 (reference)* | — | *ref.* | *ref.* | *ref.* |
| 35–40 | — | 1.428* [1.361 - 1.499] | 1.400* [1.334 - 1.469] | 1.447* [1.378 - 1.520] |
| Over 40 | — | 2.313* [2.140 - 2.500] | 2.158* [1.995 - 2.334] | 2.301* [2.125 - 2.490] |
| *White (Reference)* | — | *ref.* | *ref.* | *ref.* |
| Black | — | 1.640* [1.536 - 1.752] | 1.706* [1.596 - 1.824] | 1.744* [1.628 - 1.868] |
| South Asian | — | 0.974 [0.919 - 1.033] | 1.177* [1.109 - 1.250] | 1.098* [1.031 - 1.169] |
| Mixed ethnicity | — | 1.059 [0.957 - 1.171] | 1.116* [1.008 - 1.236] | 1.105 [0.998 - 1.224] |
| Other ethnicity | — | 0.925* [0.865 - 0.989] | 1.115* [1.041 - 1.193] | 1.116* [1.040 - 1.196] |
| Language not English | — | 0.731* [0.670 - 0.798] | 0.896* [0.821 - 0.978] | 0.938 [0.858 - 1.026] |
| BMI < 18.5 | — | — | 0.803* [0.680 - 0.948] | 0.794* [0.672 - 0.937] |
| *BMI 18.5–24.9 (reference)* | — | — | *ref.* | *ref.* |
| BMI 25.0–29.9 | — | — | 1.475* [1.392 - 1.562] | 1.475* [1.393 - 1.563] |
| BMI > 30.0 | — | — | 2.781* [2.641 - 2.929] | 2.771* [2.630 - 2.919] |
| Ever Smoker | — | — | 0.865* [0.815 - 0.919] | 0.845* [0.795 - 0.899] |
| Folic acid during pregnancy | — | — | 1.034 [0.967 - 1.107] | 1.085* [1.007 - 1.169] |
| Contacts attended percentage | — | — | 1.000 [1.000 - 1.001] | 1.001* [1.001 - 1.002] |
| Endocrine/Metabolic Diseases | — | — | 1.600* [1.505 - 1.700] | 1.625* [1.529 - 1.727] |
| Mental Disorders | — | — | 1.082* [1.027 - 1.141] | 1.082* [1.026 - 1.141] |
| Nervous System Diseases | — | — | 1.090* [1.008 - 1.179] | 1.100* [1.017 - 1.190] |
| Circulatory Diseases | — | — | 2.378* [2.230 - 2.535] | 2.393* [2.245 - 2.552] |
| Respiratory Diseases | — | — | 1.167* [1.106 - 1.231] | 1.167* [1.105 - 1.231] |
| Gastrointestinal Diseases | — | — | 1.096* [1.043 - 1.151] | 1.110* [1.057 - 1.166] |
| Musculoskeletal Diseases | — | — | 1.151* [1.072 - 1.237] | 1.152* [1.072 - 1.238] |
| Malformations/Abnormalities | — | — | 1.122 [0.942 - 1.337] | 1.012 [0.848 - 1.207] |
| AIC | 140,004.57 | 139,219.86 | 135,651.94 | 132,394.31 |
| N | 1,027,707 | 1,027,707 | 1,027,707 | 1,027,707 |
| Positive cases | 13,112 | 13,112 | 13,112 | 13,112 |

Note: The first three columns display results from models that include a single random effect for the baby's birth year, while the final column additionally includes a random effect for the hospital site where labor and delivery took place. Socioeconomic deprivation is measured as a continuous score ranging from 0.0 (least deprived) to 10.0 (most deprived); the aOR represents the change in odds per one-unit increase in score. Reference categories are shown in italics. \* = $p < 0.05$ . — indicates the variable was not included in that model.

### Socioeconomic deprivation

Increasing socioeconomic deprivation was associated with higher odds of early-onset pre-eclampsia across all models. In the confounder-adjusted model (Model 2), each one-point increase in deprivation score was associated with a 3.4% increase in odds of early-onset pre-eclampsia (aOR 1.034, 95% CI 1.027 to 1.041). Additional adjustment for theorized mediators attenuated the association modestly (aOR 1.023, 95% CI 1.017 to 1.030), suggesting that part of the deprivation association operates through individual-level health and behavioral pathways. Including hospital site as an additional random effect further attenuated the association (aOR 1.016, 95% CI 1.009 to 1.023). This pattern is consistent with systematic variation between maternity care sites contributing to observed socioeconomic inequalities in early-onset pre-eclampsia risk.

Expressed in absolute terms–with the fully adjusted model, hospital site excluded–among nulliparous women with otherwise reference-level characteristics, the predicted risk of early-onset pre-eclampsia increased from 5.0 per 1,000 pregnancies in the least deprived neighborhoods to 6.2 per 1,000 pregnancies in the most deprived neighborhoods, corresponding to an adjusted risk difference of 1.3 per 1,000 pregnancies.

Secondary analyses among multiparous women demonstrated a similar but larger deprivation association across all model specifications, although attenuation following hospital-site adjustment was less pronounced. Corresponding results for multiparous women are presented in Supplementary Table 4 (Appendix D). Analyses of late-onset pre-eclampsia showed substantially weaker deprivation associations after adjustment, particularly among nulliparous women, where effect sizes approached unity. These findings are reported in Supplementary Tables 5-6 (Appendix E).

### Covariates

Figure 4 summarizes the adjusted odds ratios from the primary nulliparous models, with and without hospital-site adjustment. Among nulliparous women in the fully adjusted model, elevated BMI was among the strongest predictors of early-onset pre-eclampsia, with obese women demonstrating nearly threefold higher odds relative to women with BMI in the healthy-weight range. Circulatory disease, maternal age over 40 years, Black ethnicity, and endocrine/metabolic disease were also strongly associated with increased risk.

**Figure 4.**
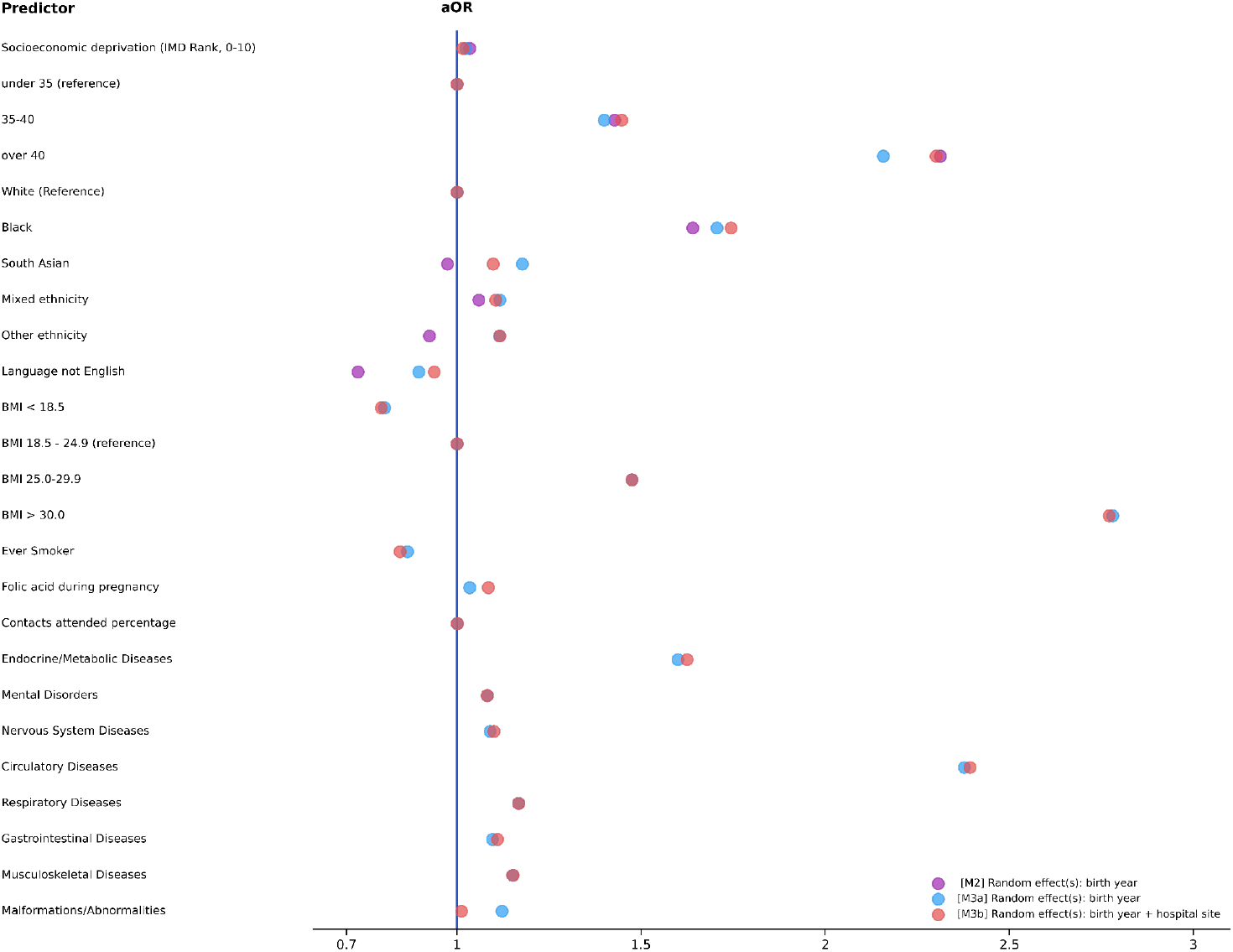
Forest plot of adjusted Odds Ratios for early-onset pre-eclampsia for nulliparous women, in Model 2 excluding the hospital-site random effect, and in Model 3 both excluding and including the hospital site random effect.

Several additional comorbidity categories demonstrated smaller but statistically significant associations with early-onset disease. Smoking was associated with reduced odds among nulliparous women, while folic acid supplementation and antenatal contact attendance showed minimal associations after adjustment.

Associations for maternal age and Black ethnicity increased slightly after inclusion of hospital-site random effects, whereas estimates for BMI and most comorbidities remained comparatively stable. Among multiparous women, the estimated association for South Asian ethnicity attenuated following hospital-site adjustment, while the effects of prior pre-eclampsia, BMI, and circulatory disease remained largely unchanged.

Among multiparous women, prior pre-eclampsia was by far the strongest predictor of recurrence, with adjusted odds ratios substantially exceeding those of all other covariates. Full regression results for multiparous women are shown in Supplementary Table 4 (Appendix D).

### Robustness Checks & Sensitivity Analyses

As a robustness check, all primary analyses were repeated using a dichotomized deprivation variable classifying women living in the most deprived 30% of neighborhoods as deprived. Results were highly consistent with those obtained using the continuous deprivation measure, although the continuous specification provided greater granularity and retained substantially more information across the deprivation distribution. Results from the dichotomized analyses are presented in Supplementary Tables 7-10. Sensitivity analyses across alternative deprivation thresholds are presented in Supplementary Figures 3-4. These investigations demonstrated stable effect estimates across alternative specifications, both with and without hospital-site random effects.

Kaplan–Meier curves (Figure 5) demonstrated minimal separation by deprivation status among nulliparous women until later gestation, whereas clearer divergence was observed among multiparous women from approximately 15 weeks onwards. Cox proportional hazards models produced estimates highly consistent with the primary logistic regression analyses across all parity groups and outcome definitions, supporting the robustness of the findings. Corresponding hazard ratio estimates are presented in Supplementary Tables 11-12 (Appendix H).

**Figure 5.**
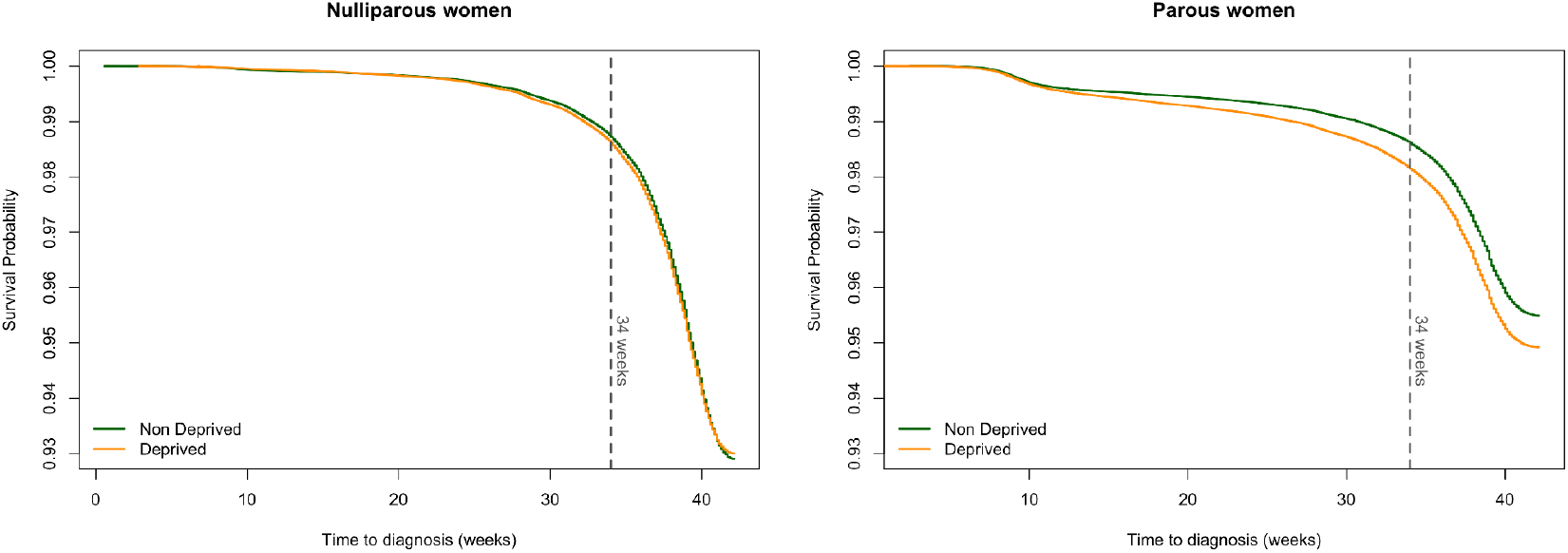
Kaplan-Meier survival curves for time to pre-eclampsia diagnosis by socioeconomic deprivation status among nulliparous and parous women.

## Discussion

### Statement of principal findings

In this national population-based cohort study of nearly two million pregnancies in England, increasing socioeconomic deprivation was associated with increased risk of early-onset pre-eclampsia, particularly among multiparous women. Associations with late-onset disease were substantially weaker after adjustment. Adjustment for theorized mediators attenuated the deprivation association modestly, while additional adjustment for hospital site further reduced the estimated effect size, suggesting that both individual-level risk factors and variation between maternity care settings contribute to observed inequalities. Elevated BMI, circulatory disease, maternal age over 40 years, Black ethnicity, and endocrine/metabolic disease were consistently associated with increased risk of early-onset pre-eclampsia. Among multiparous women, prior pre-eclampsia was the strongest predictor of recurrence.

The findings are most generalizable to singleton pregnancies receiving NHS-funded maternity care in England and may have broader relevance to other high-income healthcare systems, although effect sizes may differ due to variation in healthcare delivery structures and socioeconomic measures. Generalizability is more limited for hospital-level analyses and populations outside NHS care or with different case mix. The stronger association between deprivation and early-onset (vs. late-onset) pre-eclampsia may nonetheless extend more broadly and merits replication elsewhere.

### Strengths and weaknesses of the study

This study draws on one of the largest nationally representative maternity datasets currently available in England, with linkage across MSDS, HES, and ECDS enabling comprehensive characterization of maternal health and pregnancy outcomes. The separate analysis of early- and late-onset pre-eclampsia addresses an important limitation in the existing literature, as most previous studies have treated pre-eclampsia as a single outcome despite evidence of differing aetiologies. The use of a continuous deprivation measure derived from the full national IMD ranking allowed more granular estimation of deprivation-related risk gradients than dichotomized deprivation categories alone.

The progressive modelling framework, guided by a pre-specified DAG (Figure 1), additionally enabled examination of how adjustment for theorized mediators and hospital site altered the deprivation association. Although these nested models should not be interpreted as formal mediation analyses, they provide useful descriptive evidence regarding potential pathways through which socioeconomic inequalities may operate.

Several limitations should nonetheless be acknowledged. Deprivation was measured at neighborhood rather than individual level, introducing the possibility of ecological fallacy. BMI was estimated using maternal weight and an assumed average female height because maternal height was incompletely recorded within MSDS, and therefore should be interpreted as an approximation rather than a direct anthropometric measurement. Given its strong predictive role [23], future work using measured height would be valuable. Administrative coding practices may also vary across sites, potentially contributing to the observed hospital-level variation. Finally, residual confounding from unmeasured social, behavioral, or clinical variables cannot be excluded.

### Comparison with existing literature

Our findings are broadly consistent with prior UK evidence linking socioeconomic deprivation to adverse maternal outcomes [4, 24, 25]. However, existing studies have generally not fully addressed key dimensions relevant to clinical risk stratification: most do not distinguish between early- and late-onset pre-eclampsia, few stratify by parity, and many draw on older or regionally limited data with considerably smaller sample sizes. To our knowledge, this is the first national-scale study to separately estimate the deprivation–pre-eclampsia association by onset subtype and parity group, using a contemporary dataset spanning the entirety of England over the past five years. Prior studies have demonstrated differential risk across onset subtypes based on personal and family history, supporting the plausibility of subtype-specific socioeconomic associations [20]. The stronger deprivation association observed for early-onset disease is biologically plausible given its closer relationship to placental dysfunction and impaired uteroplacental development in the first trimester [13, 14]. Socioeconomic deprivation may impair this process through higher prevalence of chronic conditions, poorer nutrition, psychosocial stress, and delayed engagement with antenatal care, which can limit timely initiation of aspirin prophylaxis before 16 weeks’ gestation [27]. In contrast, late-onset pre-eclampsia reflects maternal cardiovascular stress later in pregnancy and may be less sensitive to early-pregnancy conditions [13,14]. The stronger association in multiparous women is consistent with the concept of biological embodiment (the cumulative physiological embedding of social disadvantage across the life course [28]) and its persistence after adjusting for prior pre-eclampsia suggests pathways beyond obstetric history.

The persistent elevation of risk among Black women after extensive adjustment is consistent with national maternal mortality surveillance and prior population-level analyses demonstrating substantial ethnic inequalities in maternal outcomes in England [5, 26]. By contrast, the association for South Asian women was attenuated after adjustment for hospital sites. This suggests that South Asians in hospital units with higher observed baseline pre-eclampsia risk are more likely to experience disparities in their outcomes.

The attenuation of deprivation associations following inclusion of hospital-site random effects is additionally consistent with wider evidence of substantial variation in maternity service quality across NHS trusts [6, 7]. While the present analysis cannot identify the specific organizational characteristics responsible for these differences, the findings suggest that inequalities in care delivery may contribute meaningfully to socioeconomic gradients in early-onset pre-eclampsia risk.

### Implications for clinicians and policymakers

The selective association between deprivation and early-onset pre-eclampsia has important clinical implications. Early-onset disease is associated with substantially greater maternal and neonatal morbidity and often requires intensified antenatal surveillance, earlier delivery, and neonatal intensive care support [13–15]. Among individual-level factors, elevated BMI demonstrated the strongest dose–response association with early-onset disease across both parity groups. Circulatory and endocrine/metabolic disease were also important predictors, supporting existing evidence linking cardiometabolic health to pre-eclampsia risk [27].

The findings additionally suggest that variation between maternity care sites may contribute independently to observed inequalities. Although NHS clinical guidelines are nationally standardized, implementation, staffing, resource availability, access to specialist services, continuity of care, and responsiveness to patient needs may differ across settings. The attenuation of deprivation associations following hospital-site adjustment is consistent with a broader “postcode lottery” in maternity outcomes, whereby women from more deprived areas may be disproportionately exposed to hospitals with higher baseline risk or fewer resources.

Importantly, these findings do not imply that hospital-level variation affects only deprived women. Rather, variation between maternity care sites may influence outcomes for all women receiving care within those settings, while socioeconomic deprivation partly shapes the distribution of patients across them.

For clinicians, these findings support heightened vigilance for early-onset pre-eclampsia among women from deprived areas, particularly those with elevated BMI or pre-existing circulatory disease. For policymakers, reducing inequalities will require both individual-level interventions and system-level efforts to address unwarranted variation, support underperforming trusts, and ensure equitable resource allocation [8].

### Unanswered questions and future research

Further work is needed to identify which organizational characteristics of maternity services drive the observed hospital-level variation. Future studies linking maternal outcomes to staffing levels, specialist access, continuity of care, guideline adherence, and Care Quality Commission ratings may help clarify these mechanisms. More detailed investigation of preventive pathways is also warranted. In particular, early-onset pre-eclampsia may be especially sensitive to timely aspirin prophylaxis and early antenatal engagement, both of which may vary systematically across socioeconomic groups. Finally, replication in other healthcare systems will be important to determine the extent to which these findings reflect features specific to NHS maternity care versus broader socioeconomic mechanisms influencing maternal health.

## Conclusion

Socioeconomic deprivation was associated with increased risk of early-onset pre-eclampsia in England among nulliparous and multiparous women. Both individual-level risk factors and variation between maternity care sites appeared to contribute to observed inequalities. Reducing these disparities will likely require coordinated efforts addressing both population health and unwarranted variation in maternity care delivery across NHS services.

## Data Availability

The study used de-identified data from the NHS Maternity Services Data Set, Hospital Episode Statistics, and the Emergency Care Data Set accessed through a data sharing agreement with NHS England. These data are subject to information governance restrictions and are not publicly available. Researchers may apply to NHS England for access through the relevant data access process, subject to approvals. Code used for data curation and analysis is available at https://github.com/ethanp274/MSDS_pre-eclampsia.

https://github.com/ethanp274/MSDS_pre-eclampsia.git

## Contributorship statement

AO, YZ, and HKM conceived and designed the study. The University of Oxford acquired funding, and managed data access. AO, YZ, and HKM provided administrative oversight and supervision, as well as ongoing monitoring of study results. CA provided clinical input and guidance on the direction of the research. MK provided input into the methodology and interpretation. EP performed data collection. EP and FCC performed data processing, literature review, coding, statistical analysis, and preparation of results. All authors contributed to the drafting of the manuscript and critically revised it for important intellectual content. AO, YZ, and HKM act as the guarantors and accept full responsibility for the work. The corresponding author attests that all listed authors meet authorship criteria and that no others meeting the criteria have been omitted. All authors have read and approved the final version of the manuscript.

## Acknowledgements

We acknowledge the support of the SDE Data Wranglers team, specifically Nickie Wareing and Angeliki Antonarou, and the MSDS team for their technical assistance. We also thank Andreas Charisiadis, Chuka Ezeoguine, Ruben Pohle, Pinelopi Stamou, and Shang Wang for their contributions to this study.

## Data sharing

The study used deidentified data from the NHS Maternity Services Data Set, Hospital Episode Statistics, and the Emergency Care Data Set accessed through a data sharing agreement with NHS England. These data are subject to information governance restrictions and are not publicly available. Researchers may apply to NHS England for access through the relevant data access process, subject to approvals. Code used for data curation and analysis is available at https://github.com/ethanp274/MSDS_pre-eclampsia.

