## Appendices for "Socioeconomic deprivation and risk of early-onset pre-eclampsia in England: a national population-based cohort study"

### Supplementary Material

#### A. Pre-processing Pipeline

The Maternity Services Data Set, and more generally the Secure Data Environment Service, provide comprehensive information on patients, hospitalization episodes, pregnancies, and associated clinical characteristics. This annex outlines the pre-processing pipeline used to construct the curated dataset underpinning the statistical analyses presented in the main text. Although the workflow diverges into multiple processing streams at several stages, these ultimately converge to produce a final, cleaned, and consolidated dataset containing all variables required for subsequent analysis. Each subsection* documents one component of the pre-processing pipeline. These components are implemented as individual Databricks notebooks, enabling a modular and reproducible workflow.

##### Stage 1: Initial Data Gathering

The master dataset was constructed from the table msds_v2_demographics_booking_and_pregnancy_all_years, which contains demographic, booking, and pregnancy information recorded in the Maternity Services Data Set. Each record is identified by the variable UniqPregId, which uniquely identifies a pregnancy and serves as the primary key for linking pregnancy-level information.

At this stage we also defined a list of variables to retain in the curated dataset for downstream analyses.

Several initial data adjustments were applied. The variable Rank_IMD_Decile_2015, which records the mother's Index of Multiple Deprivation (IMD) decile, was recoded from a string format to an integer variable. Values such as "01 – Most deprived", "02", …, "09", and "10 – Least deprived" were converted to the corresponding integers 1–10. Data types were also corrected for several numeric variables, including ageatbookingmother, gestagebooking, and previousstillbirths.

The dataset was then aggregated so that a single record was retained for each pregnancy. For each variable we applied an aggregation rule appropriate to its meaning. These included selecting the first recorded value, the last recorded value, the maximum or minimum value across records, the sum of values across records, or the collection of all observed values where multiple responses were possible.

For variables representing dates, aggregated values were converted to a standard date format.

The resulting pregnancy-level dataset was stored as msds_demographics_agg_by_uniqpregid. Only variables listed in the predefined feature list were retained for subsequent stages of the pre-processing pipeline.

##### Stage 2.1: Linkage of additional MSDS tables

The pregnancy-level dataset generated in the previous stage was extended by incorporating information from several additional tables within the Maternity Services Data Set (MSDS). For each table, selected variables were extracted and aggregated at the pregnancy level using the identifier UniqPregID.

Information from the table msds_v2_baby_activities_all_years was aggregated to derive the variables apgar_score, birth_weight, and neonatal_critical_incident.

Additional demographic and birth-related variables were obtained from msds_v2_baby_demographics_all_years. These included mother_age_at_birth, baby_age_at_death, delivery_method, baby_ethnicity, fetus_presentation, gestation_length_at_birth, birth_day_of_week, birth_am_or_pm, birth_month, birth_year, baby_phen_sex, pregnancy_outcome, neonatal_critical_care, and ld_site_id.

Maternal behavioral and physiological measures were derived from msds_v2_care_activities_all_years. Aggregation of this table produced variables describing alcohol consumption (max_alcohol_per_week, first_alcohol_per_week, last_alcohol_per_week), smoking behavior (max_cigs_per_day, first_cigs_per_day, last_cigs_per_day, smoking_status), and maternal height (mother_avg_height, mother_avg_weight) and weight (mother_first_weight, mother_last_weight).

Information on maternal critical incidents during labor was obtained from msds_v2_labor_activities_all_years. All recorded incidents were aggregated at the pregnancy level to produce the variable maternal_crit_incident.

Each aggregated table was then merged with the pregnancy-level dataset using a left join on UniqPregID, ensuring that all pregnancies present in the demographic dataset were retained. The resulting combined dataset –stored as msds_all_agg_filtered– was used as the input for subsequent pre-processing stages.

#### Stage 2.2: Mapping MSDS pregnancies to HES episodes

This stage runs in parallel to the one before. Its objective was to establish a one-to-one mapping between MSDS pregnancy records and hospital episodes recorded in Hospital Episode Statistics (HES).

Data from HES were obtained from two sources. Admitted Patient Care (APC) episodes with event type codes corresponding to labor, delivery, and birth-related episodes were retained. A separate maternity table was also extracted but not used in subsequent analyses.

The APC data were aggregated at the level of unique hospital spells, identified by the combination of patient identifier and admission date. Relevant variables were retained, and the birth year was derived from the admission date.

Reduced datasets were then prepared for matching. From HES, pregnancy-related episodes were selected based on non-missing antenatal appointment dates. From the MSDS master dataset, pregnancies with a recorded antenatal appointment date were selected. Key identifiers and appointment dates were retained from both sources.

Matching was performed by linking records with identical person identifiers, where the HES antenatal appointment date fell within three days of the MSDS antenatal appointment date. For each pregnancy, only the HES episode identifier associated with the latest labor/delivery admission date was retained.

The final mapping includes one-to-one links between MSDS pregnancies and HES episodes. Both the mapping and the aggregated HES labor/delivery episodes were saved for use in subsequent stages of the pre-processing pipeline.

#### Stage 3: Integration of HES information into the MSDS dataset

In this stage, information from Hospital Episode Statistics (HES) was integrated into the aggregated MSDS pregnancy dataset. The pregnancy-level MSDS dataset produced in Stage 2.1 was linked to the HES Admitted Patient Care (APC) data using the MSDS–HES mapping derived in Stage 2.2.

For each pregnancy, the corresponding HES record was identified via the UniqPregID–epikey linkage. Selected MSDS variables were then refined and missing values were imputed using their HES equivalents where available.

- **Birth weight (birth_weight).** Values greater than 7001 g (7 kg) were treated as implausible and set to missing. Values below 10 were assumed to be recorded in kilograms and were converted to grams. Values below 1000 g (1 kg) were also treated as implausible. Missing values were then filled using the HES variable BIRWEIT. Missingness decreased from 92.57% to 40.58%.

- **Birth outcome (pregnancy_outcome).** Only valid outcome codes corresponding to live birth or stillbirth were retained. Missing values were imputed using the HES variable BIRSTAT. Missingness decreased from 28.82% to 23.97%.

- **Gestational age at booking (gest_at_booking).** The MSDS variable was recorded in days, except for values below 50, which were assumed to be recorded in weeks and converted to days. Values corresponding to less than 10 or more than 49 weeks were treated as implausible. Missing values were imputed using the HES variable ANAGEST, which was similarly converted from weeks to days and subject to the same range restrictions. Missingness decreased from 53.08% to 46.84%.

- **Gestational age at birth (gestation_length_at_birth).** Values were converted from weeks to days when below 50 and restricted to the plausible range of 10–49 weeks. Missing values were filled using the HES variable GESTAT, with the same range restrictions applied. Missingness decreased from 29.84% to 24.84%.
- **Delivery method (delivery_method).** Invalid categorical codes were set to missing. Missing values were imputed using the HES variable DELMETH. Missingness decreased from 29.76% to 24.18%.
- **Hospital admission and discharge dates (ld_hosp_start_date, ld_disch_date).** Missing labor or delivery hospital admission dates were filled using ADMIDATE, and missing discharge dates were filled using DISDATE. Missingness decreased from 34.31% to 26.00% for admission dates and from 36.92% to 25.51% for discharge dates.
- **Hospital site identifier (ld_site_id).** Missing values were filled using the HES variable SITETRET. Missingness decreased from 44.20% to 26.17%.
- **Number of births in the pregnancy (num_births).** Missing values were filled using the HES variable NUMBABY. Missingness decreased from 26.78% to 22.84%.
- **Year and month of birth (birth_year, birth_month).** Where possible, a birth date was derived from the antenatal appointment date, gestational age at booking, and gestational age at birth:

  birth_date = antenatal_appt_date - gest_at_booking + gestation_length_at_birth

  If the derived birth date was available, the year and month components were extracted from this date. Remaining birth year missing values were then filled using the HES variable BIRYEAR. Missingness decreased from 28.65% to 23.60%.

After integration of all available HES information, the updated pregnancy-level dataset was saved as msds_joined_filtered_filled for use in the subsequent stages of the pre-processing pipeline.

#### Stage 4: Variable cleaning and recoding

In this stage, key variables in the integrated MSDS dataset were cleaned and recoded. Several categorical variables were converted into binary indicators, continuous variables were checked for implausible values, and additional derived variables were constructed.

- **Critical care indicators.** The variables neonatal_crit_incident, neonatal_critical_care, and maternal_crit_incident, originally recorded as "Y"/"N" indicators, were converted to binary variables (1/0).
- **Birth weight.** The variable birth_weight was categorized into clinically relevant groups (very low, low, normal, and high birth weight) and corresponding binary indicators were created.
- **APGAR score.** The variable apgar_score was recoded to generate indicators reflecting levels of neonatal distress, including severe or moderate distress and normal scores.
- **Birth outcome.** The categorical variable pregnancy_outcome was recoded into binary indicators distinguishing live birth and different stillbirth categories.
- **Preterm birth.** The variable gestation_length_at_birth was used to generate indicators for different categories of preterm birth (such as extremely preterm, very preterm, and moderate or late preterm).

- **Delivery method.** The variable delivery_method was recoded into binary indicators representing delivery categories such as elective or emergency cesarean section and instrumental delivery.

- **Fetal presentation.** The categorical variable fetus_presentation was recoded to binary indicators describing fetal presentation (for example cephalic, breech, or transverse).

- **Maternal age.** Maternal age at booking (age_at_booking) was restricted to plausible values between 13 and 60 years, and recoded to binary indicators for all the relevant ranges (under 35, 35 to 40, and over 40).

- **Maternal anthropometric variables.** Maternal weight variables (mother_first_weight, mother_last_weight, and mother_avg_weight) were retained and used to derive an additional variable representing weight change during pregnancy. Only maternal weight in the range the range from 35 to 200 kg were considered plausible. Maternal height values between 130 and 200 were interpreted as centimeters and converted to meters. Values below 1.3 m or above 2.0 m were treated as implausible. Body mass index was estimated as

$$mother\_bmi = \frac{mother\_avg\_weight}{female\_avg\_height^2}$$

- when weight was available, and where the national average height for women of 1.62 m was taken as the maternal height. The estimated BMI was then recoded to binary indicators according to the official NHS stratification.

- **Smoking and alcohol use.** Binary indicators were constructed to represent whether smoking or alcohol consumption had ceased by the time of booking (stopped_smoking, stopped_drinking), and whether the mother had ever partaken in any of the substances (ever_substance_use).

- **Additional binary indicators.** Several variables originally recorded as "Y"/"N" were converted to binary indicators, including abnormal_ultrasound_ind, social_factors_ind, disability_ind, and support_ind.

- **Folic acid supplementation.** The variable folic_acid was recoded into indicators describing folic acid use before pregnancy, during pregnancy, or no recorded use.

- **Sociodemographic variables.** Categorical variables describing ethnicity, parental employment, maternal language, religion, sexual orientation, and the baby's sex were recoded into sets of binary indicator variables.

- **Index of Multiple Deprivation (IMD).** Two additional variables were derived from the IMD decile: an indicator for missing IMD values and an indicator for residence in a deprived area (IMD decile ≤3).

- **Obstetric history.** Previous pregnancy statistics (such as the amount of previous live births, cesarean sections, 24 weeks losses and stillbirths) were recoded as binary indicators.

- **Contact counts.** The number of attended contacts was restricted to values between 1 and 40. The same approach was applied to the variable describing face-to-face contacts. Additional variables were derived to characterize patterns of care utilization, including a binary indicator for a higher-than-normal number of contacts (more than 20), the proportion of face-to-face contacts among all contacts, and a binary indicator for a high proportion of face-to-face contacts (greater than 33.33%).

- **Time of birth.** Binary indicators were created for each day of the week and each month using the variables birth_day_of_week and birth_month. Additional indicators were derived for the trimester of birth and for the time of day (based on birth_am_or_pm). Binary indicators for the year of birth (2018–2025) were also derived from birth_year.

- **Timing of antenatal care.** Two timing variables were constructed: days_to_first_appointment, defined as the number of days between the last menstrual period and the first antenatal appointment, and is_mother_late, an indicator for late presentation (more than 90 days). When the variable was missing, it was imputed using the first antenatal appointment date minus 16 weeks.

After selecting only the processed features to keep, the resulting dataset was retained for subsequent stages of the pre-processing pipeline as msds_processed_cleaned_vars.

#### Stage 5.1: Emergency Readmissions

This step identified for each pregnancy whether the mother experienced an emergency hospital readmission within 42 days following discharge from labor and delivery services. The dataset msds_processed_cleaned_vars produced in the previous step was loaded as the master data frame, from which a reduced table was then created containing only the variables required for this task: UniqPregID, person_id_deid, and ld_disch_date.

Emergency readmission was defined as a hospital spell beginning within 42 days of the labor or delivery discharge date in which the mother received a diagnosis related to pregnancy or postpartum complications, which corresponds to the common definition of the *puerperium*. Readmissions were identified using three national hospital data sources that record diagnostic information using different clinical coding systems.

- **Hospital Episode Statistics – Accident and Emergency (HES AE).** Emergency department attendances were loaded and aggregated by patient identifier and arrival date to obtain one record per hospital spell. All diagnosis codes recorded during each spell were collected into a list of diagnosis codes.

- **Hospital Episode Statistics – Admitted Patient Care (HES APC).** Hospital admissions were loaded and aggregated by patient identifier and admission date, again producing one record per hospital spell. Diagnoses were recorded using ICD-10 codes.

- **Emergency Care Data Set (ECDS).** Emergency care records were loaded and aggregated by patient identifier and arrival date. Diagnoses in this dataset were recorded using SNOMED CT terminology.

For each dataset, hospital spells were restricted to those occurring within 42 days after the labor or delivery discharge date. Diagnosis code description tables were then filtered to retain only codes associated with postpartum complications. Filtering was performed using keywords chosen to capture the postpartum readmission causes described in Table 2 of a previous study on postpartum readmissions (Pritchett et al., *BJOG*, 2025;132:178–88;doi:10.1111/1471-0528.17955). For each hospital spell, the list of diagnosis codes was compared with the filtered code lists, and spells containing at least one postpartum-related diagnosis were retained.

Distinct UniqPregID values from the three filtered datasets were then combined into a single set representing pregnancies with a postpartum emergency readmission.

A binary indicator, emergency_readmission, was added to the master dataset and takes the value 1 when the pregnancy identifier appears in this set, and 0 otherwise. Approximately 5.19% of pregnancies were classified as having an emergency readmission. This information was joined back into the initial complete data frame, which would later serve as the master data set for the next stage.

The resulting dataset was then saved as msds_with_readmission for use in subsequent analyses.

#### Stage 5.2: Diagnoses and Comorbidities

This step identified key maternal diagnoses and comorbidities occurring before and during the pregnancy, with a particular focus on pre-eclampsia and gestational diabetes. It used multiple national healthcare data sources, including emergency care, hospital admissions, and maternity records, and was conducted in parallel with other outcome derivations.

The dataset msds_processed_cleaned_vars was loaded into the data frame df_master.

As a preliminary step, a reduced pregnancy-level dataset was constructed for subsequent busyness analysis. The labor and delivery site identifier, start date, and end date were refined by drawing on additional MSDS sources where the primary values are missing, and invalid or unresolvable site codes were replaced with null values. The end date was further adjusted such that spells exceeding 14 days in duration were truncated to 14 days from the start date, and spells with identical start and end dates were extended by one day. The dataset was then filtered to retain only pregnancies with non-null values for the site identifier, start date, and end date, and where the start date did not exceed the end date. This reduced dataset was saved as msds_ld_date for use in the next stage.

For the identification of diagnoses and comorbidities, another reduced pregnancy-level dataset was constructed containing UniqPregID, person_id_deid, last_period_date, antenatal_appt_date, and labor and delivery dates. Where necessary, missing last_period_date values were approximated using the antenatal appointment date.

Diagnostic information was then extracted from multiple data sources (including MSDS, HES admitted patient care, HES emergency attendances, and emergency care datasets). For each source, records were linked to pregnancies using the patient identifier and classified into two time periods:

- pre-pregnancy, defined as records occurring before the last menstrual period;
- during pregnancy, defined as records occurring between the last menstrual period and the labor or delivery discharge date.

Within each period, all recorded diagnoses and comorbidities were aggregated at the pregnancy level. Clinical codes from different coding systems (ICD-10 and SNOMED CT) were mapped to a selection of clinical concepts using standard reference descriptions.

Binary indicators were then derived to capture the presence of key conditions. In particular:

- preeclampsia: indicating whether any diagnosis consistent with pre-eclampsia or related hypertensive disorders of pregnancy was recorded before or during pregnancy;
- gestational_diabetes: indicating whether gestational diabetes was recorded before or during pregnancy.

Additional broader diagnostic groupings exploring medical history and conditions were constructed for later analyses but are not central to the primary outcomes.

All diagnosis-derived variables were combined into a single pregnancy-level dataset, all_datasets_diagnoses, with one record per pregnancy. This dataset is used in subsequent analyses.

#### Stage 6: Final data assembly, cohort restriction, and imputation

In this step, the outputs of the previous data processing steps were combined to produce the final analytical dataset. This includes cleaned MSDS variables, emergency readmission indicators, and diagnosis-derived variables.

The datasets msds_with_readmission, all_datasets_diagnoses, and msds_ld_date were merged at the pregnancy level using the unique pregnancy identifier (UniqPregID). All variables required for subsequent analyses were retained.

Additional variables were derived at this stage, including:

- ld_hosp_length_stay, defined as the duration between labor or delivery admission and discharge;
- previous_pregnancy, a binary indicator of whether the mother had a recorded prior pregnancy.

The merged dataset initially contained roughly 4 million pregnancy records. A series of restrictions was then applied to define the analytical cohort:

- exclusion of multiple births;
- restriction to pregnancies with recorded birth year from 2021 onward;
- restriction to records with non-missing deprivation index (IMD);
- consistency between birth year and labor or delivery discharge year (difference of at most one year);

Records with a missing hospital identifier were also filled with a placeholder value.

Intermediate diagnostic variables used during processing were removed, and the resulting dataset was saved as msds_diag_busy_filtered_final.

To prepare for imputation, the dataset was partitioned into subgroups defined by combinations of birth year and IMD decile, and imputation was performed separately within each subgroup to preserve the underlying distribution of variables across time and socioeconomic strata. Only binary variables were imputed; categorical variables, dates, identifiers, and selected structural variables were excluded and retained as reference. Missing values were filled using $k$-nearest neighbors imputation with $k = 5$, where $k$ denotes the number of most similar complete observations used to estimate each missing value.

Finally, all imputed subsets were recombined to produce the complete analytical dataset, saved as msds_diag_busy_filtered_final_imputed, which is used in subsequent modeling.

#### Stage 7.1: Pre-eclampsia indicators

This step identified indicators of pre-eclampsia during pregnancy, based on diagnoses, clinical findings, and observations. Three key components were considered: hypertension, proteinuria, and placental growth factor (PlGF).

The dataset msds_diag_busy_filtered_final_imputed was loaded and restricted to the first recorded pregnancy per mother. Relevant identifiers and pregnancy dates (last_period_date and ld_hosp_disch_date) were retained.

Diagnostic and clinical information was obtained from multiple sources. Findings and observations were extracted from MSDS, while diagnoses were sourced from MSDS, hospital admissions, and emergency care records. MSDS diagnoses were restricted to pregnancy-related entries.

Relevant clinical codes were identified using keyword-based searches across coding systems (including SNOMED CT and ICD-10), supplemented by manual selection where appropriate. No specific codes were identified for abnormalities in placental growth factor nor evidence of testing. The full list of manual codes and key-words can be found in Supplementary Table 1.

Supplementary Table 1. Diagnosis code and keyword definitions

| **Condition** | **SNOMED-CT codes** | **ICD-10 codes** | **Keywords** |
|---|---|---|---|
| Pre-eclampsia | 15938005, 15938005, 29738008, 71701000119105, 72022006, 198992004, 169560008, 398254007, 427889009, 48194001, 69909000, 111479006 | O14, O14.0, O14.1, O14.2, O14.9, O11, O15, O15.0, O15.1, O15.2, O15.9, O10, O13, O16 | — |
| Hypertension | *Diastolic blood pressure*<br>271650006, 1091811000000102, 716632005<br>*Systolic blood pressure*<br>72313002, 271649006, 716579001, 72313002 | — | Single: "hypertension",<br>Sets: ["high", "blood", "pressure"], ["elevated", "blood", "pressure"] |
| Proteinuria | — | — | Single: "proteinuria" |

Supplementary Table 1. Diagnosis code and keyword definitions

*Note:* This table shows the diagnosis phenotyping rules used in the analysis code. Clinical codes and keyword patterns are rendered in monospaced text to distinguish them from narrative descriptions. Section labels inside cells are explanatory comments; a dash indicates no entry was used for that condition. SNOMED-CT and ICD-10 codes and keywords—both manually selected—for pre-eclampsia, hypertension and proteinuria diagnosis.

For each indicator, data sources were processed separately. Records were first filtered using keyword-based searches to retain only entries corresponding to the indicator of interest. These filtered records were then linked to the reduced pregnancy-level dataset (containing identifiers and pregnancy dates) and restricted to events occurring between the start and end of pregnancy. Records from all sources were subsequently combined and grouped by pregnancy identifier and event date, so that each occurrence of the indicator were retained. Counts of occurrences during pregnancy were then derived for each indicator: Hypertension_count, Proteinuria_count, and PlGF_count.

Binary indicators were also derived:

- Hypertension_2x: at least two recorded hypertension events during pregnancy;
- Proteinuria_1x: at least one recorded proteinuria event;
- PlGF_test: evidence of any placental growth factor test.

The resulting dataset was saved as msds_diag_busy_filtered_final_imputed_reduced for subsequent stages of pre-processing.

#### Stage 7.2: Provisional pre-eclampsia diagnosis with timing

In parallel to the identification of pre-eclampsia indicators, this stage derived a provisional pre-eclampsia diagnosis incorporating temporal information, enabling classification into early and late onset.

The dataset obtained in the previous step (Stage 6) was used as the master data frame. Although a pre-eclampsia indicator was previously constructed (Stage 5.2), the derivation was repeated here to incorporate diagnosis dates required for temporal classification.

Diagnoses were identified from the Maternity Services Data Set (MSDS), the Emergency Care Data Set (ECDS), and Hospital Episode Statistics admitted patient care data (HES APC). Hospital Episode Statistics accident and emergency data (HES AE) were initially considered but yielded no relevant diagnoses and were therefore excluded.

Pre-eclampsia-related codes were selected from ICD-10 and SNOMED CT.

For each source, data were first loaded and linked to the pregnancy-level dataset to retain relevant entries. Records were then restricted to events occurring between the start (last_period_date) and end (ld_hosp_disch_date) of pregnancy, and filtered to include only diagnoses containing at least one pre-eclampsia-related code. Diagnoses were subsequently grouped across sources by pregnancy identifier, retaining the earliest recorded diagnosis.

A binary variable (Preeclampsia_during_this_pregnancy) was constructed to indicate the presence of a diagnosis during pregnancy. Among positive cases, time to diagnosis was calculated as the interval between diagnosis date and estimated pregnancy start—set at 40 weeks before the estimated delivery date. This interval was used to classify pre-eclampsia as early or late onset. Early onset pre-eclampsia (Preeclampsia_early_onset) is defined as a diagnosis occurring before 34 weeks' gestation and was set to 1 in such cases and 0 otherwise. Late onset pre-eclampsia (Preeclampsia_late_onset) is defined as a diagnosis occurring at or after 34 weeks' gestation; it was set to 1 for such cases, 0 for pregnancies without a diagnosis, and null for pregnancies classified as early onset.

The resulting dataset was saved for subsequent processing as msds_diag_busy_filtered_final_imputed_reduced_timed.

#### Stage 8: Final pre-eclampsia diagnosis

This final pre-processing stage defined the main outcomes by combining the provisional pre-eclampsia diagnosis (stage 7.2) with indicator-based evidence derived in stage 7.1.

The dataset obtained in the previous steps was merged—with a left join that preserved the filtering at the beginning of stage 7.1—and used as the master data frame. An additional indicator (Possible_preeclampsia_diagnosis_AND1x_or_Hyp2x) was constructed from clinical findings: pregnancies with at least one recorded instance of both hypertension and proteinuria, or with at least two counts of hypertension, were considered to have a possible pre-eclampsia diagnosis.

The primary outcome variable (Preeclampsia_during_this_pregnancy) was then updated to identify pregnancies with both a recorded diagnosis and supporting clinical evidence; that is, cases where both the provisional diagnosis and the indicator-based definition are present.

Early and late onset classifications were subsequently aligned with the updated outcome definition. Early onset pre-eclampsia (Preeclampsia_early_onset) is defined as a diagnosis occurring before 34 weeks' gestation and was set to 0 for pregnancies without a confirmed diagnosis. Late onset pre-eclampsia (Preeclampsia_late_onset) is defined as a diagnosis occurring at or after 34 weeks' gestation; it was set to 0 for pregnancies without a confirmed diagnosis and to missing for pregnancies classified as early onset, so that early cases were not included in the late onset group.

At this stage, the Index of Multiple Deprivation (IMD) rank corresponding to the mother's Lower Layer Super Output Area (LSOA) was linked to the dataset, drawn from the English Indices of Multiple Deprivation 2019. In the original ranking, 1 denotes the most deprived neighborhood and 32,844 the least deprived. A scaled continuous variable (IMD_Rank_scaled), ranging from 0 to 10, was derived as a complementary deprivation measure alongside the binary indicator deprived_reg; the scale was reversed relative to the original ranking, such that 0 indicates least deprivation and 10 indicates greatest deprivation, so that higher values correspond intuitively to higher pre-eclampsia risk in the regression analysis.

Before the pre-processing phase is concluded, two additional modifications were applied to the dataset. The variable indicating whether the mother had ever smoked was redefined in accordance with NHS guidelines, and a new variable was derived representing the proportion of scheduled antenatal contacts that were attended.

The population was then stratified into two mutually exclusive groups for use in subsequent analyses:

- **Group 1:** nulliparous women with no prior diagnosis of pre-eclampsia or gestational diabetes mellitus.
- **Group 2:** multiparous women.

The resulting dataset—saved as msds_diag_busy_filtered_final_imputed_reduced_timed_ind— represents the final version of the master data set, to be used in the subsequent regression analysis.

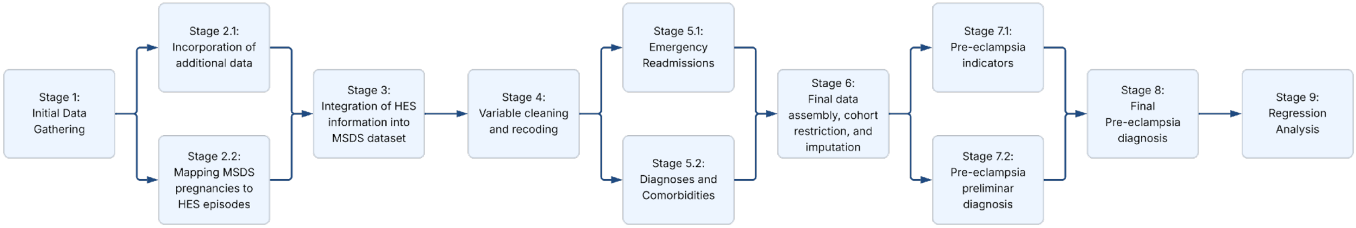


**Supplementary Figure 1.** Full processing pipeline

### B. Variable Dictionary

Supplementary Table 2. Variable dictionary

| Variables | Description | Type(s) |
|---|---|---|
| **Regressors** | | |
| Socioeconomic deprivation | The IMD (Index of Multiple Deprivation) ranks every lower-layer super output area (LSOA) in England from 1 (most deprived) to 32,844 (least deprived). Each woman's socioeconomic deprivation is represented as a continuous score derived by inverting and rescaling this rank, such that 0.0 corresponds to the least deprived neighborhood and 10.0 to the most deprived; higher values indicate greater deprivation. | Continuous |
| Maternal age | Maternal age at booking, categorized into risk-based groups: < 35 (reference), 35-40, and >40 years. | Binary |
| Ethnicity | The self-reported maternal ethnicity recorded at booking, recorded into one of the following categories: White (reference), Black, South Asian, Mixed or Other. | Binary |
| Language not English | Indicates whether the mother speaks English or not. | Binary |
| Maternal BMI | Maternal body mass index categories based on NHS definitions: underweight (<18.5), healthy weight (18.5-24.9, reference), overweight (25.0-29.9), and obesity (≥30). | Binary |
| Ever Smoker | Indicates whether the mother ever smoked tobacco. | Binary |
| Folic acid during pregnancy | Reports whether there was any intake of folic acid during the pregnancy. | Binary |
| Contacts attended percentage | Percentage of attended contacts, calculated over the total amount of scheduled contacts. | Continuous |
| Previous pre-eclampsia | Indicates whether the mother was diagnosed with pre-eclampsia in a previous pregnancy. | Binary |
| Previous gestational DM | Indicates whether the mother was diagnosed with gestational diabetes mellitus in a previous pregnancy. | Binary |
| Endocrine/Metabolic Diseases | Indicates whether the mother has any endocrine or metabolic diseases. | Binary |
| Mental Disorders | Indicates whether the mother has ever been diagnosed with any mental disorder. | Binary |
| Nervous System Diseases | Indicates whether the mother has ever been diagnosed with any nervous system disease. | Binary |
| Circulatory Diseases | Indicates whether the mother has any circulatory diseases. | Binary |
| Respiratory Diseases | Indicates whether the mother has ever been diagnosed with any respiratory disease. | Binary |
| Gastrointestinal Diseases | Indicates whether the mother has ever been diagnosed with any gastrointestinal disease. | Binary |
| Musculoskeletal Diseases | Indicates whether the mother has any musculoskeletal diseases. | Binary |
| Malformations/Abnormalities | Indicates whether the mother has reported any malformations or abnormalities. | Binary |
| **Random effects** | | |
| Birth Year | Report the birth year of the baby corresponding to the pregnancy. | Categorical |
| Hospital Site ID | Report the identifier of the hospital where the mother gave birth, if the birth occurred at an NHS site. | Categorical |
| **Outcomes** | | |
| Pre-eclampsia | Indicates whether the mother has been diagnosed with pre-eclampsia throughout the pregnancy. | Binary |
| Early Pre-eclampsia (< 34 weeks) | Indicates whether the mother has been diagnosed with pre-eclampsia early in the pregnancy, i.e. before 34 weeks of gestation. | Binary |
| Late Pre-eclampsia (≥ 34 weeks) | Indicates whether the mother has been diagnosed with pre-eclampsia early in the pregnancy, i.e. from 34 weeks of gestation onwards. | Binary |

*Note:* This dictionary lists the variables used in the regression models, together with their short descriptions and data types. It does not include every variable available in the raw dataset.

### C. Data missingness

Supplementary Table 3. Variable missingness (pre- and post-imputation)

| Variable | Pre-imputation | Post-imputation |
|---|---|---|
| **Regressors** | | |
| Socioeconomic deprivation (IMD Rank, 0-10) | — | 0.00% |
| Maternal Age | | |
| Under 35 (reference) | 0.00% | 0.00% |
| 35–40 | 0.00% | 0.00% |
| Over 40 | 0.00% | 0.00% |
| Ethnicity | | |
| White (Reference) | 3.70% | 0.00% |
| Black | 3.70% | 0.00% |
| South Asian | 3.70% | 0.00% |
| Mixed ethnicity | 3.70% | 0.00% |
| Other ethnicity | 3.70% | 0.00% |
| Preferred Language not English | 40.27% | 0.00% |
| Maternal BMI | | |
| BMI < 18.5 | 33.41% | 0.00% |
| BMI 18.5–24.9 (reference) | 33.41% | 0.00% |
| BMI 25.0–29.9 | 33.41% | 0.00% |
| BMI > 30.0 | 33.41% | 0.00% |
| Ever Smoker* | 0.00% | 0.00% |
| Folic acid during pregnancy | 16.42% | 0.00% |
| Contacts attended percentage* | 17.42% | 0.00% |
| Medical History | | |
| Previous pre-eclampsia | 0.96% | 0.00% |
| Previous gestational DM | 0.96% | 0.00% |
| Endocrine/Metabolic Diseases | 0.96% | 0.00% |
| Mental Disorders | 0.96% | 0.00% |
| Nervous System Diseases | 0.96% | 0.00% |
| Circulatory Diseases | 0.96% | 0.00% |
| Respiratory Diseases | 0.96% | 0.00% |
| Gastrointestinal Diseases | 0.96% | 0.00% |
| Musculoskeletal Diseases | 0.96% | 0.00% |
| Malformations/Abnormalities | 0.96% | 0.00% |
| **Random Effects** | | |
| Birth Year | 0.00% | 0.00% |
| Hospital Site ID | 0.00% | 0.00% |
| **Outcomes** | | |
| Pre-eclampsia* | — | 0.00% |
| Early Pre-eclampsia (< 34 weeks)* | — | 0.00% |
| Late Pre-eclampsia (≥ 34 weeks)* | — | 1.43% |

*Note:* The post-imputation variable names are kept as the row labels. A star (*) marks rows where the pre-imputation column uses an alternative pre-imputation variable ("Ever Smoking" instead of "Ever Smoker", "Contacts attended" instead of "Contacts attended percentage") or simply a null value, where the variable did not exist before imputation.

### D. Regression results for early-onset pre-eclampsia among multiparous women

Supplementary Table 4. Regression results for early-onset pre-eclampsia in multiparous women with continuous IMD — Models 1, 2 and 3 with birth-year random effect only, and Model 3 with birth-year + hospital-site random effects

| Regressor | Model 1 | Model 2 | Model 3 | |
|---|---|---|---|---|
| | **Early PE**<br>**Hosp excluded**<br>aOR [95% CI] | **Early PE**<br>**Hosp excluded**<br>aOR [95% CI] | **Early PE**<br>**Hosp excluded**<br>aOR [95% CI] | **Early PE**<br>**Hosp included**<br>aOR [95% CI] |
| *Intercept (base odds)* | 0.012* [0.010 - 0.016] | 0.010* [0.008 - 0.012] | 0.003* [0.002 - 0.003] | 0.003* [0.002 - 0.003] |
| Socioeconomic deprivation | 1.061* [1.055 - 1.067] | 1.063* [1.057 - 1.069] | 1.046* [1.040 - 1.053] | 1.041* [1.034 - 1.049] |
| *Under 35 (reference)* | – | *ref.* | *ref.* | *ref.* |
| 35–40 | – | 1.542* [1.486 - 1.600] | 1.628* [1.567 - 1.692] | 1.652* [1.589 - 1.717] |
| Over 40 | – | 2.362* [2.237 - 2.493] | 2.462* [2.325 - 2.606] | 2.523* [2.382 - 2.673] |
| *White (Reference)* | – | *ref.* | *ref.* | *ref.* |
| Black | – | 1.774* [1.681 - 1.871] | 1.889* [1.784 - 2.000] | 1.942* [1.829 - 2.063] |
| South Asian | – | 1.130* [1.073 - 1.191] | 1.372* [1.298 - 1.450] | 1.260* [1.187 - 1.337] |
| Mixed ethnicity | – | 1.009 [0.912 - 1.117] | 1.104 [0.995 - 1.226] | 1.100 [0.990 - 1.222] |
| Other ethnicity | – | 0.838* [0.784 - 0.896] | 1.095* [1.022 - 1.174] | 1.090* [1.014 - 1.170] |
| Language not English | – | 0.832* [0.772 - 0.897] | 0.978 [0.905 - 1.056] | 1.051 [0.972 - 1.136] |
| BMI < 18.5 | – | – | 0.778* [0.635 - 0.953] | 0.782* [0.638 - 0.957] |
| *BMI 18.5–24.9 (reference)* | – | – | *ref.* | *ref.* |
| BMI 25.0–29.9 | – | – | 1.309* [1.233 - 1.390] | 1.297* [1.221 - 1.377] |
| BMI > 30.0 | – | – | 2.313* [2.192 - 2.440] | 2.268* [2.149 - 2.394] |
| Ever Smoker | – | – | 1.100* [1.045 - 1.158] | 1.047 [0.993 - 1.104] |
| Folic acid during pregnancy | – | – | 0.987 [0.934 - 1.043] | 1.053 [0.992 - 1.116] |
| Contacts attended percentage | – | – | 1.003* [1.002 - 1.003] | 1.003* [1.002 - 1.004] |
| Previous pre-eclampsia | – | – | 9.543* [9.201 - 9.899] | 9.396* [9.055 - 9.749] |
| Previous gestational DM | – | – | 1.174* [1.108 - 1.244] | 1.201* [1.132 - 1.273] |
| Endocrine/Metabolic Diseases | – | – | 1.150* [1.105 - 1.198] | 1.149* [1.103 - 1.197] |
| Mental Disorders | – | – | 1.022 [0.979 - 1.067] | 1.011 [0.968 - 1.055] |
| Nervous System Diseases | – | – | 1.215* [1.144 - 1.290] | 1.218* [1.146 - 1.294] |
| Circulatory Diseases | – | – | 2.447* [2.328 - 2.572] | 2.487* [2.365 - 2.615] |
| Respiratory Diseases | – | – | 1.105* [1.058 - 1.155] | 1.107* [1.059 - 1.156] |
| Gastrointestinal Diseases | – | – | 1.096* [1.051 - 1.144] | 1.100* [1.054 - 1.148] |
| Musculoskeletal Diseases | – | – | 1.053 [0.998 - 1.111] | 1.036 [0.982 - 1.094] |
| Malformations/Abnormalities | – | – | 1.212* [1.034 - 1.420] | 1.198* [1.019 - 1.410] |
| AIC | 148,789.27 | 147,115.89 | 129,129.86 | 125,565.52 |
| N | 940,505 | 940,505 | 940,505 | 940,505 |
| Positive cases | 14,526 | 14,526 | 14,526 | 14,526 |

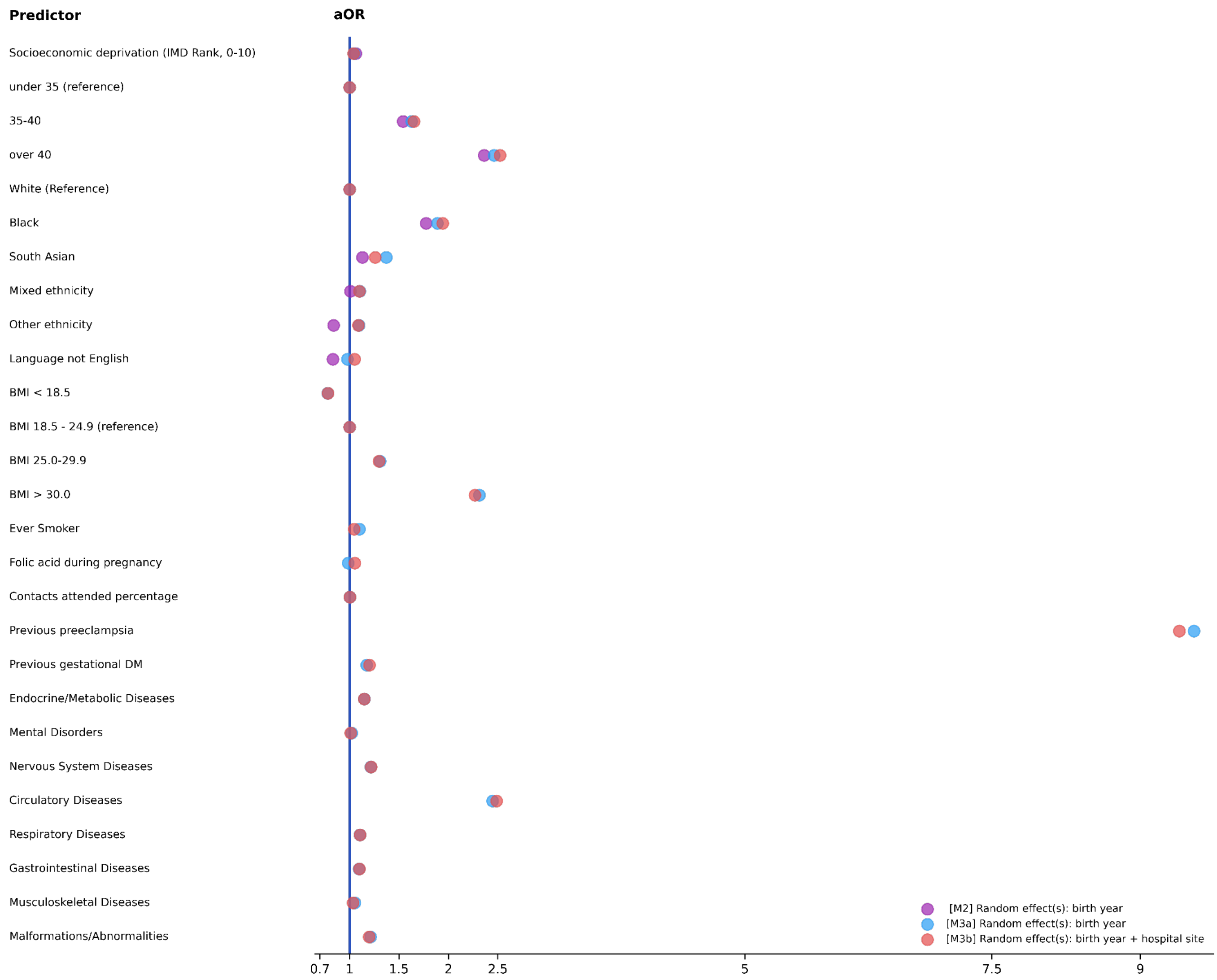


**Supplementary Figure 2.** Forest plot of adjusted Odds Ratios for early-onset pre-eclampsia for parous women, in Model 2 excluding the hospital-site random effect, and in Model 3 both excluding and including the hospital site random effect.

### E. Regression results for late-onset pre-eclampsia

Supplementary Table 5. Regression results for late-onset pre-eclampsia in nulliparous women with continuous IMD — Models 1, 2 and 3 with birth-year random effect only, and Model 3 with birth-year + hospital-site random effects

| Regressor | Model 1 | Model 2 | Model 3 | |
|---|---|---|---|---|
| | **Late PE** **Hosp excluded** aOR [95% CI] | **Late PE** **Hosp excluded** aOR [95% CI] | **Late PE** **Hosp excluded** aOR [95% CI] | **Late PE** **Hosp included** aOR [95% CI] |
| *Intercept (base odds)* | 0.065* [0.056 - 0.076] | 0.066* [0.057 - 0.078] | 0.030* [0.026 - 0.035] | 0.037* [0.035 - 0.039] |
| Socioeconomic deprivation | 0.999 [0.996 - 1.002] | 1.006* [1.003 - 1.009] | 1.005* [1.002 - 1.008] | 1.005* [1.002 - 1.008] |
| *Under 35 (reference)* | – | *ref.* | *ref.* | *ref.* |
| 35–40 | – | 1.056* [1.029 - 1.082] | 1.045* [1.019 - 1.072] | 1.069* [1.042 - 1.097] |
| Over 40 | – | 1.106* [1.051 - 1.164] | 1.082* [1.028 - 1.139] | 1.142* [1.084 - 1.202] |
| *White (Reference)* | – | *ref.* | *ref.* | *ref.* |
| Black | – | 1.140* [1.099 - 1.183] | 1.111* [1.070 - 1.153] | 1.213* [1.168 - 1.261] |
| South Asian | – | 0.750* [0.727 - 0.772] | 0.825* [0.800 - 0.850] | 0.827* [0.800 - 0.854] |
| Mixed ethnicity | – | 0.742* [0.702 - 0.784] | 0.762* [0.720 - 0.805] | 0.783* [0.740 - 0.829] |
| Other ethnicity | – | 0.727* [0.702 - 0.752] | 0.797* [0.770 - 0.826] | 0.824* [0.794 - 0.854] |
| Language not English | – | 0.902* [0.866 - 0.940] | 0.963 [0.925 - 1.004] | 1.008 [0.967 - 1.051] |
| BMI < 18.5 | – | – | 0.754* [0.700 - 0.813] | 0.765* [0.710 - 0.824] |
| *BMI 18.5–24.9 (reference)* | – | – | *ref.* | *ref.* |
| BMI 25.0–29.9 | – | – | 1.372* [1.337 - 1.407] | 1.356* [1.322 - 1.391] |
| BMI > 30.0 | – | – | 2.212* [2.160 - 2.266] | 2.170* [2.118 - 2.222] |
| Ever Smoker | – | – | 0.892* [0.866 - 0.918] | 0.874* [0.848 - 0.900] |
| Folic acid during pregnancy | – | – | 0.970 [0.939 - 1.001] | 1.064* [1.027 - 1.103] |
| Contacts attended percentage | – | – | 1.005* [1.005 - 1.006] | 1.003* [1.002 - 1.003] |
| Endocrine/Metabolic Diseases | – | – | 1.123* [1.082 - 1.166] | 1.144* [1.102 - 1.188] |
| Mental Disorders | – | – | 0.862* [0.837 - 0.887] | 0.898* [0.873 - 0.925] |
| Nervous System Diseases | – | – | 1.058* [1.011 - 1.107] | 1.060* [1.013 - 1.110] |
| Circulatory Diseases | – | – | 1.212* [1.160 - 1.266] | 1.213* [1.161 - 1.267] |
| Respiratory Diseases | – | – | 1.038* [1.009 - 1.069] | 1.034* [1.004 - 1.065] |
| Gastrointestinal Diseases | – | – | 0.974* [0.949 - 0.999] | 0.972* [0.947 - 0.997] |
| Musculoskeletal Diseases | – | – | 0.985 [0.944 - 1.027] | 0.984 [0.944 - 1.026] |
| Malformations/Abnormalities | – | – | 0.750* [0.663 - 0.848] | 0.897 [0.793 - 1.015] |
| AIC | 451,468.74 | 450,494.21 | 443,347.9 | 431,204.9 |
| N | 1,014,481 | 1,014,481 | 1,014,481 | 1,014,481 |
| Positive cases | 59,437 | 59,437 | 59,437 | 59,437 |

*Note:* The denominator for the late pre-eclampsia model (N=1,014,481) reflects the exclusion of 13,226 individuals from the main-text cohort (N=1,027,707). This total comprises the 13,112 cases of early-onset pre-eclampsia and 114 cases where the onset timing could not be classified due to a missing reference date.

Supplementary Table 6. Regression results for late-onset pre-eclampsia in multiparous women with continuous IMD — Models 1, 2 and 3 with birth-year random effect only, and Model 3 with birth-year + hospital-site random effects

| Regressor | Model 1 | Model 2 | Model 3 | |
|---|---|---|---|---|
| | **Late PE**<br>**Hosp excluded**<br>aOR [95% CI] | **Late PE**<br>**Hosp excluded**<br>aOR [95% CI] | **Late PE**<br>**Hosp excluded**<br>aOR [95% CI] | **Late PE**<br>**Hosp included**<br>aOR [95% CI] |
| *Intercept (base odds)* | 0.035* [0.029 - 0.041] | 0.030* [0.025 - 0.035] | 0.012* [0.010 - 0.014] | 0.014* [0.013 - 0.015] |
| Socioeconomic deprivation | 1.010* [1.006 - 1.014] | 1.014* [1.009 - 1.018] | 1.013* [1.008 - 1.017] | 1.017* [1.013 - 1.022] |
| *Under 35 (reference)* | – | *ref.* | *ref.* | *ref.* |
| 35–40 | – | 1.338* [1.304 - 1.374] | 1.372* [1.336 - 1.409] | 1.391* [1.354 - 1.429] |
| Over 40 | – | 1.870* [1.794 - 1.949] | 1.924* [1.845 - 2.007] | 1.975* [1.893 - 2.061] |
| *White (Reference)* | – | *ref.* | *ref.* | *ref.* |
| Black | – | 1.373* [1.316 - 1.433] | 1.305* [1.249 - 1.364] | 1.394* [1.331 - 1.460] |
| South Asian | – | 0.938* [0.903 - 0.975] | 1.012 [0.972 - 1.054] | 1.023 [0.979 - 1.068] |
| Mixed ethnicity | – | 0.876* [0.813 - 0.945] | 0.911* [0.844 - 0.984] | 0.934 [0.864 - 1.008] |
| Other ethnicity | – | 0.847* [0.809 - 0.887] | 0.943* [0.900 - 0.989] | 0.937* [0.893 - 0.984] |
| Language not English | – | 1.055* [1.002 - 1.110] | 1.114* [1.057 - 1.174] | 1.083* [1.026 - 1.143] |
| BMI < 18.5 | – | – | 0.900 [0.795 - 1.020] | 0.884 [0.779 - 1.003] |
| *BMI 18.5–24.9 (reference)* | – | – | *ref.* | *ref.* |
| BMI 25.0–29.9 | – | – | 1.339* [1.288 - 1.393] | 1.348* [1.296 - 1.403] |
| BMI > 30.0 | – | – | 2.199* [2.121 - 2.279] | 2.205* [2.125 - 2.287] |
| Ever Smoker | – | – | 0.883* [0.849 - 0.918] | 0.914* [0.878 - 0.952] |
| Folic acid during pregnancy | – | – | 0.981 [0.944 - 1.020] | 1.020 [0.978 - 1.064] |
| Contacts attended percentage | – | – | 1.003* [1.003 - 1.004] | 1.001* [1.001 - 1.002] |
| Previous pre-eclampsia | – | – | 6.086* [5.912 - 6.265] | 6.162* [5.984 - 6.345] |
| Previous gestational DM | – | – | 1.100* [1.050 - 1.152] | 1.123* [1.071 - 1.176] |
| Endocrine/Metabolic Diseases | – | – | 1.020 [0.989 - 1.051] | 1.049* [1.017 - 1.081] |
| Mental Disorders | – | – | 0.869* [0.842 - 0.896] | 0.878* [0.850 - 0.906] |
| Nervous System Diseases | – | – | 1.059* [1.008 - 1.113] | 1.063* [1.012 - 1.118] |
| Circulatory Diseases | – | – | 1.408* [1.346 - 1.472] | 1.432* [1.368 - 1.498] |
| Respiratory Diseases | – | – | 0.978 [0.946 - 1.011] | 0.973 [0.941 - 1.006] |
| Gastrointestinal Diseases | – | – | 1.008 [0.977 - 1.040] | 0.999 [0.968 - 1.031] |
| Musculoskeletal Diseases | – | – | 0.980 [0.939 - 1.023] | 0.969 [0.929 - 1.012] |
| Malformations/Abnormalities | – | – | 0.881 [0.767 - 1.013] | 1.045 [0.907 - 1.205] |
| AIC | 264,258.2 | 262,867.19 | 246,617.73 | 240,125.52 |
| N | 925,934 | 925,934 | 925,934 | 925,934 |
| Positive cases | 30,042 | 30,042 | 30,042 | 30,042 |

*Note:* The denominator for the late pre-eclampsia model (N=925,934) reflects the exclusion of all early-onset pre-eclampsia cases and the remaining 45 cases where the onset timing could not be classified due to a missing reference date. Together with the 114 unclassifiable cases excluded from the nulliparous cohort, these constitute the 159 unclassifiable cases described in the main text.

### F[1]. Binary socioeconomic deprivation variable robustness check

Supplementary Table 7. Regression results for early-onset pre-eclampsia in nulliparous women with dichotomous IMD — Models 1, 2 and 3 with birth-year random effect only, and Model 3 with birth-year + hospital-site random effects

| Regressor | Model 1 | Model 2 | Model 3 | |
|---|---|---|---|---|
| | **Early PE**<br>**Hosp excluded**<br>aOR [95% CI] | **Early PE**<br>**Hosp excluded**<br>aOR [95% CI] | **Early PE**<br>**Hosp excluded**<br>aOR [95% CI] | **Early PE**<br>**Hosp included**<br>aOR [95% CI] |
| *Intercept (base odds)* | 0.013* [0.011 - 0.016] | 0.012* [0.010 - 0.014] | 0.006* [0.005 - 0.007] | 0.006* [0.005 - 0.006] |
| Socioeconomic deprivation | 1.085* [1.048 - 1.124] | 1.102* [1.062 - 1.143] | 1.056* [1.018 - 1.096] | 1.011 [0.971 - 1.052] |
| *Under 35 (reference)* | – | *ref.* | *ref.* | *ref.* |
| 35–40 | – | 1.409* [1.343 - 1.478] | 1.387* [1.322 - 1.456] | 1.438* [1.370 - 1.510] |
| Over 40 | – | 2.278* [2.107 - 2.462] | 2.135* [1.974 - 2.309] | 2.283* [2.110 - 2.472] |
| *White (Reference)* | – | *ref.* | *ref.* | *ref.* |
| Black | – | 1.686* [1.578 - 1.802] | 1.744* [1.632 - 1.865] | 1.780* [1.662 - 1.907] |
| South Asian | – | 0.993 [0.936 - 1.103] | 1.197* [1.128 - 1.271] | 1.113* [1.045 - 1.185] |
| Mixed ethnicity | – | 1.069 [0.966 - 1.183] | 1.125* [1.016 - 1.245] | 1.115* [1.007 - 1.235] |
| Other ethnicity | – | 0.937 [0.876 - 1.002] | 1.128* [1.054 - 1.208] | 1.129* [1.053 - 1.211] |
| Language not English | – | 0.750* [0.687 - 0.819] | 0.915* [0.839 - 0.998] | 0.957 [0.875 - 1.046] |
| BMI < 18.5 | – | – | 0.813* [0.688 - 0.959] | 0.801* [0.679 - 0.946] |
| *BMI 18.5–24.9 (reference)* | – | – | *ref.* | *ref.* |
| BMI 25.0–29.9 | – | – | 1.475* [1.392 - 1.563] | 1.475* [1.393 - 1.563] |
| BMI > 30.0 | – | – | 2.795* [2.655 - 2.944] | 2.781* [2.639 - 2.929] |
| Ever Smoker | – | – | 0.874* [0.823 - 0.928] | 0.852* [0.801 - 0.906] |
| Folic acid during pregnancy | – | – | 1.019 [0.952 - 1.090] | 1.070 [0.993 - 1.153] |
| Contacts attended percentage | – | – | 1.000 [1.000 - 1.001] | 1.001* [1.001 - 1.002] |
| Endocrine/Metabolic Diseases | – | – | 1.603* [1.509 - 1.704] | 1.628* [1.532 - 1.730] |
| Mental Disorders | – | – | 1.090* [1.034 - 1.149] | 1.087* [1.031 - 1.146] |
| Nervous System Diseases | – | – | 1.091* [1.009 - 1.180] | 1.100* [1.018 - 1.190] |
| Circulatory Diseases | – | – | 2.380* [2.233 - 2.538] | 2.394* [2.245 - 2.553] |
| Respiratory Diseases | – | – | 1.171* [1.110 - 1.236] | 1.170* [1.109 - 1.235] |
| Gastrointestinal Diseases | – | – | 1.096* [1.044 - 1.152] | 1.111* [1.057 - 1.167] |
| Musculoskeletal Diseases | – | – | 1.152* [1.072 - 1.237] | 1.153* [1.073 - 1.239] |
| Malformations/Abnormalities | – | – | 1.116 [0.937 - 1.330] | 1.007 [0.844 - 1.201] |
| AIC | 140,075.44 | 139,309.24 | 135,701.1 | 132,420.21 |

[1] The sample size for this dichotomous analysis includes an additional 139 cases compared to the continuous analysis models (comprising 88 cases in the early-onset analysis and 51 cases in the late-onset analysis). These 139 cases—which correspond to the specific exclusions detailed in the participant flow diagram (Figure 2)—lacked an exact Lower-layer Super Output Area (LSOA) match, meaning their continuous deprivation score could not be derived. However, because this missing data represents a negligible fraction of the total cohort (nearly 2 million pregnancies), these cases were retained in the dichotomous models to maximize sample size.

| | | | | |
|---|---|---|---|---|
| N | 1,027,795 | 1,027,795 | 1,027,795 | 1,027,795 |
| Positive cases | 13,113 | 13,113 | 13,113 | 13,113 |

Supplementary Table 8. Regression results for early-onset pre-eclampsia in multiparous women with dichotomous IMD — Models 1, 2 and 3 with birth-year random effect only, and Model 3 with birth-year + hospital-site random effects

| | **Model 1** | **Model 2** | **Model 3** | |
|---|---|---|---|---|
| **Regressor** | **Early PE Hosp excluded** aOR [95% CI] | **Early PE Hosp excluded** aOR [95% CI] | **Early PE Hosp excluded** aOR [95% CI] | **Early PE Hosp included** aOR [95% CI] |
| *Intercept (base odds)* | 0.016* [0.012 - 0.020] | 0.012* [0.010 - 0.015] | 0.003* [0.003 - 0.004] | 0.003* [0.003 - 0.004] |
| Socioeconomic deprivation | 1.327* [1.284 - 1.371] | 1.315* [1.271 - 1.361] | 1.228* [1.184 - 1.273] | 1.193* [1.148 - 1.241] |
| *Under 35 (reference)* | – | *ref.* | *ref.* | *ref.* |
| 35–40 | – | 1.518* [1.463 - 1.575] | 1.613* [1.552 - 1.676] | 1.641* [1.579 - 1.706] |
| Over 40 | – | 2.323* [2.200 - 2.452] | 2.436* [2.302 - 2.579] | 2.507* [2.367 - 2.655] |
| *White (Reference)* | – | *ref.* | *ref.* | *ref.* |
| Black | – | 1.810* [1.715 - 1.909] | 1.919* [1.813 - 2.032] | 1.972* [1.857 - 2.095] |
| South Asian | – | 1.147* [1.088 - 1.208] | 1.393* [1.318 - 1.472] | 1.274* [1.201 - 1.352] |
| Mixed ethnicity | – | 1.015 [0.917 - 1.123] | 1.108 [0.999 - 1.230] | 1.106 [0.996 - 1.229] |
| Other ethnicity | – | 0.847* [0.792 - 0.905] | 1.108* [1.034 - 1.187] | 1.102* [1.026 - 1.184] |
| Language not English | – | 0.846* [0.785 - 0.912] | 0.991 [0.918 - 1.070] | 1.064 [0.984 - 1.150] |
| BMI < 18.5 | – | – | 0.786* [0.642 - 0.962] | 0.788* [0.644 - 0.965] |
| *BMI 18.5–24.9 (reference)* | – | – | *ref.* | *ref.* |
| BMI 25.0–29.9 | – | – | 1.313* [1.237 - 1.394] | 1.300* [1.224 - 1.380] |
| BMI > 30.0 | – | – | 2.336* [2.214 - 2.464] | 2.285* [2.164 - 2.412] |
| Ever Smoker | – | – | 1.113* [1.057 - 1.172] | 1.055* [1.001 - 1.113] |
| Folic acid during pregnancy | – | – | 0.975 [0.922 - 1.030] | 1.042 [0.982 - 1.105] |
| Contacts attended percentage | – | – | 1.003* [1.002 - 1.003] | 1.003* [1.002 - 1.004] |
| Previous pre-eclampsia | – | – | 9.531* [9.189 - 9.885] | 9.384* [9.044 - 9.736] |
| Previous gestational DM | – | – | 1.175* [1.109 - 1.245] | 1.204* [1.135 - 1.276] |
| Endocrine/Metabolic Diseases | – | – | 1.152* [1.107 - 1.200] | 1.150* [1.104 - 1.198] |
| Mental Disorders | – | – | 1.032 [0.989 - 1.077] | 1.018 [0.975 - 1.063] |
| Nervous System Diseases | – | – | 1.215* [1.144 - 1.291] | 1.218* [1.146 - 1.294] |
| Circulatory Diseases | – | – | 2.453* [2.334 - 2.578] | 2.491* [2.369 - 2.619] |
| Respiratory Diseases | – | – | 1.107* [1.060 - 1.157] | 1.108* [1.060 - 1.158] |
| Gastrointestinal Diseases | – | – | 1.057* [1.002 - 1.115] | 1.100* [1.055 - 1.148] |
| Musculoskeletal Diseases | – | – | 1.057* [1.002 - 1.115] | 1.039 [0.984 - 1.097] |
| Malformations/Abnormalities | – | – | 1.207* [1.030 - 1.414] | 1.196* [1.017 - 1.406] |

| | | | | |
|---|---|---|---|---|
| AIC | 148,944.99 | 147,303.4 | 129,224.51 | 125,631.79 |
| N | 940,556 | 940,556 | 940,556 | 940,556 |
| Positive cases | 14,530 | 14,530 | 14,530 | 14,530 |

Supplementary Table 9. Regression results for late-onset pre-eclampsia in nulliparous women with dichotomous IMD — Models 1, 2 and 3 with birth-year random effect only, and Model 3 with birth-year + hospital-site random effects

| | **Model 1** | **Model 2** | **Model 3** | |
|---|---|---|---|---|
| **Regressor** | **Late PE**<br>**Hosp excluded**<br>aOR [95% CI] | **Late PE**<br>**Hosp excluded**<br>aOR [95% CI] | **Late PE**<br>**Hosp excluded**<br>aOR [95% CI] | **Late PE**<br>**Hosp included**<br>aOR [95% CI] |
| *Intercept (base odds)* | 0.066* [0.057 - 0.076] | 0.069* [0.059 - 0.080] | 0.031* [0.027 - 0.036] | 0.038* [0.036 - 0.040] |
| Socioeconomic deprivation | 0.962* [0.946 - 0.979] | 1.000 [0.982 - 1.018] | 0.998 [0.980 - 1.016] | 1.006 [0.986 - 1.026] |
| *Under 35 (reference)* | – | *ref.* | *ref.* | *ref.* |
| 35–40 | – | 1.051* [1.025 - 1.078] | 1.042* [1.016 - 1.069] | 1.067* [1.040 - 1.095] |
| Over 40 | – | 1.100* [1.046 - 1.158] | 1.078* [1.024 - 1.135] | 1.139* [1.082 - 1.200] |
| *White (Reference)* | – | *ref.* | *ref.* | *ref.* |
| Black | – | 1.153* [1.111 - 1.196] | 1.121* [1.080 - 1.164] | 1.220* [1.174 - 1.268] |
| South Asian | – | 0.755* [0.733 - 0.778] | 0.830* [0.805 - 0.856] | 0.830* [0.803 - 0.857] |
| Mixed ethnicity | – | 0.745* [0.705 - 0.788] | 0.764* [0.723 - 0.808] | 0.785* [0.742 - 0.831] |
| Other ethnicity | – | 0.731* [0.706 - 0.756] | 0.801* [0.774 - 0.830] | 0.827* [0.797 - 0.857] |
| Language not English | – | 0.912* [0.875 - 0.950] | 0.972 [0.933 - 1.013] | 1.014 [0.973 - 1.057] |
| BMI < 18.5 | – | – | 0.757* [0.703 - 0.816] | 0.767* [0.712 - 0.828] |
| *BMI 18.5–24.9 (reference)* | – | – | *ref.* | *ref.* |
| BMI 25.0–29.9 | – | – | 1.372* [1.337 - 1.407] | 1.356* [1.322 - 1.391] |
| BMI > 30.0 | – | – | 2.216* [2.164 - 2.269] | 2.172* [2.121 - 2.225] |
| Ever Smoker | – | – | 0.895* [0.870 - 0.921] | 0.876* [0.850 - 0.902] |
| Folic acid during pregnancy | – | – | 0.964* [0.933 - 0.995] | 1.060* [1.022 - 1.098] |
| Contacts attended percentage | – | – | 1.005* [1.005 - 1.006] | 1.003* [1.002 - 1.003] |
| Endocrine/Metabolic Diseases | – | – | 1.123* [1.082 - 1.166] | 1.144* [1.102 - 1.188] |
| Mental Disorders | – | – | 0.864* [0.839 - 0.889] | 0.900* [0.874 - 0.926] |
| Nervous System Diseases | – | – | 1.058* [1.011 - 1.107] | 1.060* [1.013 - 1.110] |
| Circulatory Diseases | – | – | 1.212* [1.160 - 1.267] | 1.213* [1.161 - 1.268] |
| Respiratory Diseases | – | – | 1.039* [1.010 - 1.070] | 1.035* [1.005 - 1.065] |
| Gastrointestinal Diseases | – | – | 0.974 [0.949 - 1.000] | 0.972* [0.947 - 0.998] |
| Musculoskeletal Diseases | – | – | 0.985 [0.945 - 1.028] | 0.985 [0.944 - 1.027] |
| Malformations/Abnormalities | – | – | 0.749* [0.662 - 0.846] | 0.896 [0.792 - 1.014] |
| AIC | 451524.78 | 450585.26 | 443429.14 | 431280.2 |

| | | | | |
|---|---|---|---|---|
| N | 1,014,568 | 1,014,568 | 1,014,568 | 1,014,568 |
| Positive cases | 59,444 | 59,444 | 59,444 | 59,444 |

Supplementary Table 10. Regression results for late-onset pre-eclampsia in multiparous women with dichotomous IMD — Models 1, 2 and 3 with birth-year random effect only, and Model 3 with birth-year + hospital-site random effects

| Regressor | **Model 1** | **Model 2** | **Model 3** | |
|---|---|---|---|---|
| | **Late PE**<br>**Hosp excluded**<br>aOR [95% CI] | **Late PE**<br>**Hosp excluded**<br>aOR [95% CI] | **Late PE**<br>**Hosp excluded**<br>aOR [95% CI] | **Late PE**<br>**Hosp included**<br>aOR [95% CI] |
| *Intercept (base odds)* | 0.036* [0.030 - 0.043] | 0.031* [0.027 - 0.037] | 0.013* [0.011 - 0.015] | 0.015* [0.014 - 0.016] |
| Socioeconomic deprivation | 1.032* [1.008 - 1.056] | 1.038* [1.013 - 1.063] | 1.035* [1.009 - 1.061] | 1.057* [1.028 - 1.087] |
| *Under 35 (reference)* | – | *ref.* | *ref.* | *ref.* |
| 35–40 | – | 1.330* [1.296 - 1.365] | 1.366* [1.330 - 1.403] | 1.385* [1.348 - 1.423] |
| Over 40 | – | 1.858* [1.783 - 1.936] | 1.915* [1.836 - 1.998] | 1.967* [1.894 - 2.052] |
| *White (Reference)* | – | *ref.* | *ref.* | *ref.* |
| Black | – | 1.391* [1.333 - 1.451] | 1.322* [1.265 - 1.381] | 1.411* [1.348 - 1.478] |
| South Asian | – | 0.946* [0.910 - 0.983] | 1.022 [0.982 - 1.064] | 1.031 [0.987 - 1.076] |
| Mixed ethnicity | – | 0.880* [0.817 - 0.949] | 0.915* [0.848 - 0.988] | 0.938 [0.869 - 1.013] |
| Other ethnicity | – | 0.852* [0.814 - 0.892] | 0.950* [0.907 - 0.996] | 0.945* [0.900 - 0.992] |
| Language not English | – | 1.066* [1.012 - 1.122] | 1.125* [1.068 - 1.185] | 1.094* [1.037 - 1.154] |
| BMI < 18.5 | – | – | 0.905 [0.799 - 1.026] | 0.889 [0.784 - 1.008] |
| *BMI 18.5–24.9 (reference)* | – | – | *ref.* | *ref.* |
| BMI 25.0–29.9 | – | – | 1.341* [1.289 - 1.394] | 1.350* [1.297 - 1.404] |
| BMI > 30.0 | – | – | 2.209* [2.131 - 2.289] | 2.214* [2.135 - 2.297] |
| Ever Smoker | – | – | 0.889* [0.855 - 0.924] | 0.920* [0.884 - 0.958] |
| Folic acid during pregnancy | – | – | 0.974 [0.937 - 1.013] | 1.013 [0.971 - 1.056] |
| Contacts attended percentage | – | – | 1.003* [1.003 - 1.004] | 1.001* [1.001 - 1.002] |
| Previous pre-eclampsia | – | – | 6.079* [5.905 - 6.257] | 6.153* [5.976 - 6.336] |
| Previous gestational DM | – | – | 1.099* [1.050 - 1.152] | 1.123* [1.072 - 1.177] |
| Endocrine/Metabolic Diseases | – | – | 1.020 [0.990 - 1.051] | 1.049* [1.018 - 1.082] |
| Mental Disorders | – | – | 0.873* [0.846 - 0.901] | 0.882* [0.855 - 0.911] |
| Nervous System Diseases | – | – | 1.059* [1.008 - 1.113] | 1.064* [1.012 - 1.118] |
| Circulatory Diseases | – | – | 1.409* [1.347 - 1.474] | 1.433* [1.369 - 1.499] |
| Respiratory Diseases | – | – | 0.979 [0.947 - 1.012] | 0.974 [0.942 - 1.007] |
| Gastrointestinal Diseases | – | – | 1.009 [0.978 - 1.041] | 0.999 [0.968 - 1.032] |
| Musculoskeletal Diseases | – | – | 0.981 [0.941 - 1.024] | 0.970 [0.930 - 1.013] |
| Malformations/Abnormalities | – | – | 0.880 [0.766 - 1.011] | 1.044 [0.906 - 1.203] |

| | | | | |
|---|---|---|---|---|
| AIC | 264280.64 | 262903.05 | 246644.98 | 240163.71 |
| N | 925,981 | 925,981 | 925,981 | 925,981 |
| Positive cases | 30,042 | 30,042 | 30,042 | 30,042 |

#### G. Socioeconomic deprivation threshold sensitivity analysis

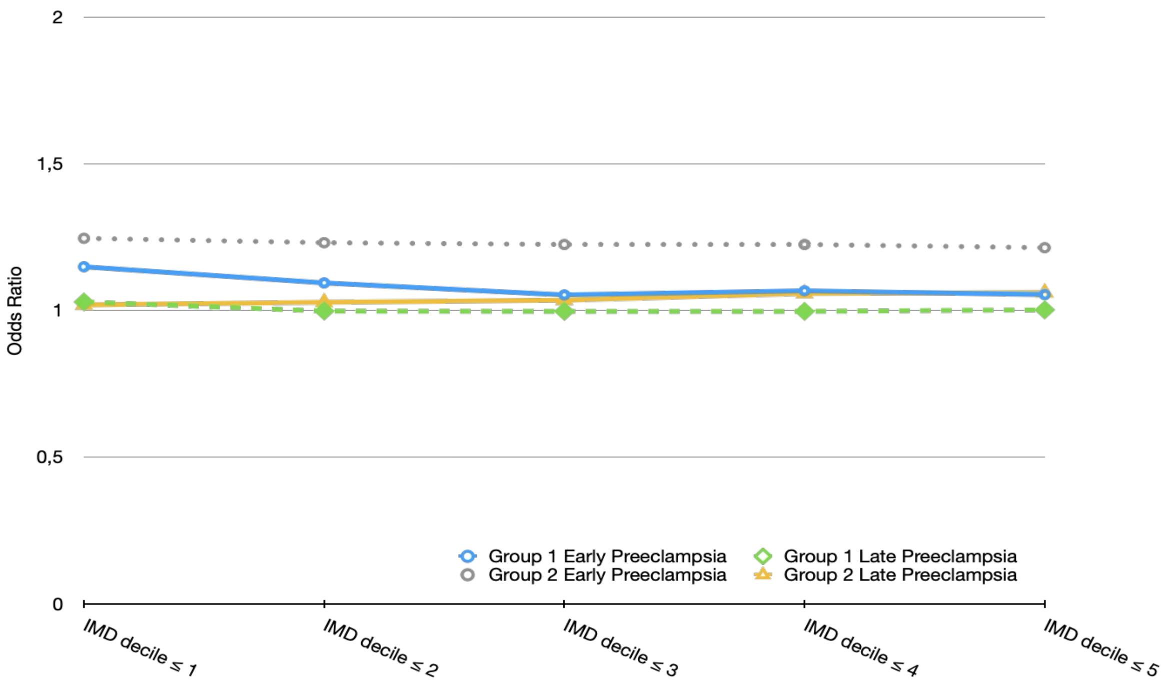


**Supplementary Figure 3.** Robustness check of the association between socioeconomic deprivation and pre-eclampsia across alternative IMD thresholds. Adjusted odds ratios for socioeconomic deprivation are shown for five deprivation definitions (IMD deciles ≤1 to ≤5) from the fully adjusted model (Model 3), separately for early pre-eclampsia (<34 weeks) and late pre-eclampsia (≥34 weeks), in Groups 1 and 2. The plot shows estimates from the model including birth year as a random effect.

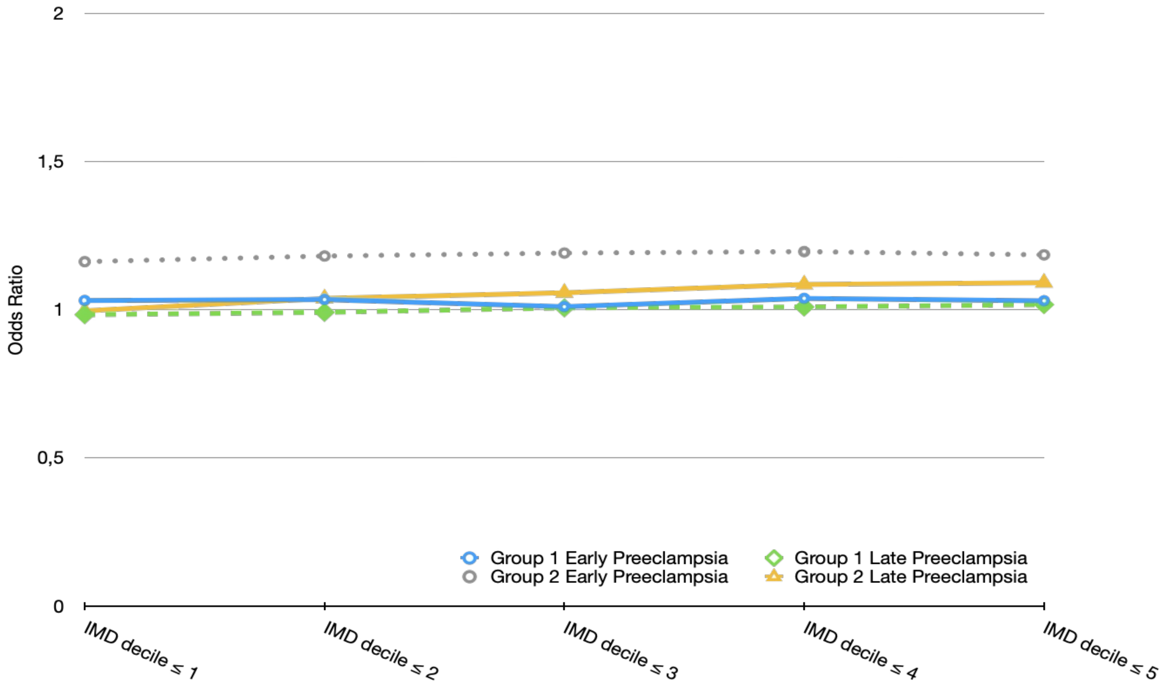


**Supplementary Figure 4.** Robustness check of the association between socioeconomic deprivation and pre-eclampsia across alternative IMD thresholds. Adjusted odds ratios for socioeconomic deprivation are shown for five deprivation definitions (IMD deciles ≤1 to ≤5) from the fully adjusted model (Model 3), separately for early pre-eclampsia (<34 weeks) and late pre-eclampsia (≥34 weeks), in Groups 1 and 2. The plot shows estimates from the model including birth year and hospital site as random effects.

### H. Cox Proportional Hazard Models

Supplementary Table 11. Cox proportional hazards results for nulliparous women — Hospital effects excluded

| | Model 1 | | Model 2 | | Model 3 | |
|---|---|---|---|---|---|---|
| **Regressor** | **Early PE (<34 wks)** | **Late PE (≥34 wks)** | **Early PE (<34 wks)** | **Late PE (≥34 wks)** | **Early PE (<34 wks)** | **Late PE (≥34 wks)** |
| Socioeconomic deprivation | 1.085* [1.047, 1.124] | 0.965* [0.949, 0.981] | 1.092* [1.053, 1.133] | 0.994 [0.977, 1.012] | 1.053* [1.016, 1.092] | 0.994 [0.976, 1.011] |
| *under 35 (reference)* | – | – | *ref.* | *ref.* | *ref.* | *ref.* |
| 35–40 | – | – | 1.405* [1.340, 1.474] | 1.048* [1.023, 1.074] | 1.392* [1.327, 1.460] | 1.042* [1.017, 1.068] |
| over 40 | – | – | 2.261* [2.094, 2.442] | 1.102* [1.049, 1.157] | 2.137* [1.979, 2.308] | 1.085* [1.032, 1.139] |
| *White (Reference)* | – | – | *ref.* | *ref.* | *ref.* | *ref.* |
| Black | – | – | 1.678* [1.572, 1.791] | 1.149* [1.109, 1.191] | 1.750* [1.639, 1.869] | 1.128* [1.088, 1.169] |
| South Asian | – | – | 0.987 [0.932, 1.046] | 0.750* [0.729, 0.772] | 1.164* [1.098, 1.234] | 0.816* [0.792, 0.840] |
| Mixed ethnicity | – | – | 1.073 [0.971, 1.187] | 0.752* [0.712, 0.794] | 1.117* [1.010, 1.236] | 0.769* [0.728, 0.812] |
| Other ethnicity | – | – | 0.933* [0.873, 0.997] | 0.735* [0.711, 0.760] | 1.103* [1.031, 1.180] | 0.800* [0.774, 0.828] |
| Language not English | – | – | 0.792* [0.726, 0.865] | 1.017 [0.978, 1.058] | 0.912* [0.835, 0.996] | 1.040 [1.000, 1.081] |
| BMI < 18.5 | – | – | – | – | 0.636* [0.532, 0.760] | 0.682* [0.632, 0.736] |
| *BMI 18.5–24.9 (reference)* | – | – | – | – | *ref.* | *ref.* |
| BMI 25.0–29.9 | – | – | – | – | 1.231* [1.174, 1.290] | 1.230* [1.205, 1.257] |
| BMI > 30.0 | – | – | – | – | 2.097* [2.014, 2.184] | 1.823* [1.789, 1.859] |
| Ever Smoker | – | – | – | – | 0.871* [0.821, 0.925] | 0.894* [0.870, 0.919] |
| Folic acid during pregnancy | – | – | – | – | 1.018 [0.952, 1.088] | 0.970 [0.940, 1.000] |
| Contacts attended percentage | – | – | – | – | 1.000 [0.999, 1.000] | 1.005* [1.005, 1.005] |
| Endocrine/Metabolic Diseases | – | – | – | – | 1.661* [1.565, 1.762] | 1.173* [1.132, 1.216] |
| Mental Disorders | – | – | – | – | 1.101* [1.045, 1.159] | 0.886* [0.862, 0.911] |
| Nervous System Diseases | – | – | – | – | 1.096* [1.015, 1.183] | 1.065* [1.020, 1.112] |
| Circulatory Diseases | – | – | – | – | 2.361* [2.218, 2.514] | 1.216* [1.167, 1.268] |
| Respiratory Diseases | – | – | – | – | 1.183* [1.122, 1.247] | 1.055* [1.026, 1.085] |

Supplementary Table 11. Cox proportional hazards results for nulliparous women — Hospital effects excluded

| | Model 1 | | Model 2 | | Model 3 | |
|---|---|---|---|---|---|---|
| Gastrointestinal Diseases | – | – | – | – | 1.100*<br>[1.048, 1.154] | 0.983<br>[0.959, 1.008] |
| Musculoskeletal Diseases | – | – | – | – | 1.156*<br>[1.078, 1.240] | 0.999<br>[0.960, 1.040] |
| Malformations/Abnormalities | – | – | – | – | 1.128<br>[0.951, 1.337] | 0.790*<br>[0.703, 0.889] |

*Note:* Values are hazard ratios with 95% confidence intervals in brackets. Early PE = Early-onset pre-eclampsia (<34 weeks); Late PE = Late-onset pre-eclampsia (≥34 weeks). '—' indicates the variable was not included in that model. * = p<0.05

Supplementary Table 12. Cox proportional hazards results for multiparous women — Hospital effects excluded

| | Model 1 | | Model 2 | | Model 3 | |
|---|---|---|---|---|---|---|
| **Regressor** | **Early PE (<34 wks)** | **Late PE (≥34 wks)** | **Early PE (<34 wks)** | **Late PE (≥34 wks)** | **Early PE (<34 wks)** | **Late PE (≥34 wks)** |
| Socioeconomic deprivation | 1.323*<br>[1.281, 1.367] | 1.032*<br>[1.009, 1.056] | 1.302*<br>[1.258, 1.346] | 1.038*<br>[1.014, 1.063] | 1.218*<br>[1.177, 1.261] | 1.043*<br>[1.018, 1.068] |
| *under 35 (reference)* | – | – | *ref.* | *ref.* | *ref.* | *ref.* |
| 35–40 | – | – | 1.511*<br>[1.456, 1.567] | 1.324*<br>[1.290, 1.359] | 1.584*<br>[1.527, 1.643] | 1.353*<br>[1.319, 1.389] |
| over 40 | – | – | 2.294*<br>[2.175, 2.420] | 1.839*<br>[1.766, 1.914] | 2.321*<br>[2.199, 2.449] | 1.873*<br>[1.800, 1.951] |
| *White (Reference)* | – | – | *ref.* | *ref.* | *ref.* | *ref.* |
| Black | – | – | 1.799*<br>[1.707, 1.896] | 1.383*<br>[1.327, 1.442] | 1.864*<br>[1.766, 1.967] | 1.325*<br>[1.271, 1.382] |
| South Asian | – | – | 1.122*<br>[1.066, 1.182] | 0.942*<br>[0.907, 0.978] | 1.343*<br>[1.274, 1.417] | 1.003<br>[0.965, 1.043] |
| Mixed ethnicity | – | – | 1.020<br>[0.922, 1.127] | 0.883*<br>[0.820, 0.951] | 1.105<br>[0.999, 1.222] | 0.917*<br>[0.852, 0.988] |
| Other ethnicity | – | – | 0.843*<br>[0.789, 0.901] | 0.854*<br>[0.817, 0.894] | 1.087*<br>[1.017, 1.163] | 0.945*<br>[0.903, 0.989] |
| Language not English | – | – | 0.955<br>[0.889, 1.027] | 1.111*<br>[1.057, 1.168] | 1.042<br>[0.968, 1.120] | 1.128*<br>[1.073, 1.186] |
| BMI < 18.5 | – | – | – | – | 0.613*<br>[0.488, 0.769] | 0.660*<br>[0.573, 0.760] |
| *BMI 18.5–24.9 (reference)* | – | – | – | – | *ref.* | *ref.* |
| BMI 25.0–29.9 | – | – | – | – | 1.147*<br>[1.094, 1.203] | 1.123*<br>[1.088, 1.158] |
| BMI > 30.0 | – | – | – | – | 1.843*<br>[1.769, 1.919] | 1.684*<br>[1.638, 1.731] |
| Ever Smoker | – | – | – | – | 1.104*<br>[1.051, 1.160] | 0.889*<br>[0.856, 0.923] |
| Folic acid during pregnancy | – | – | – | – | – | 0.987 |

Supplementary Table 12. Cox proportional hazards results for multiparous women — Hospital effects excluded

| Regressor | Model 1 | | Model 2 | | Model 3 | |
|---|---|---|---|---|---|---|
| | **Early PE (<34 wks)** | **Late PE (≥34 wks)** | **Early PE (<34 wks)** | **Late PE (≥34 wks)** | **Early PE (<34 wks)** | **Late PE (≥34 wks)** |
| | | | | | | [0.951, 1.025] |
| Contacts attended percentage | – | – | – | – | 1.002* [1.002, 1.003] | 1.003* [1.003, 1.003] |
| Previous pre-eclampsia | – | – | – | – | 8.937* [8.631, 9.255] | 5.656* [5.506, 5.811] |
| Previous gestational DM | – | – | – | – | 1.164* [1.102, 1.229] | 1.111* [1.063, 1.161] |
| Endocrine/Metabolic Diseases | – | – | – | – | 1.188* [1.143, 1.235] | 1.062* [1.032, 1.093] |
| Mental Disorders | – | – | – | – | 1.037 [0.995, 1.080] | 0.882* [0.856, 0.910] |
| Nervous System Diseases | – | – | – | – | 1.202* [1.136, 1.273] | 1.060* [1.011, 1.112] |
| Circulatory Diseases | – | – | – | – | 2.317* [2.211, 2.427] | 1.390* [1.332, 1.451] |
| Respiratory Diseases | – | – | – | – | 1.106* [1.060, 1.153] | 0.985 [0.954, 1.017] |
| Gastrointestinal Diseases | – | – | – | – | 1.091* [1.048, 1.136] | 1.012 [0.982, 1.043] |
| Musculoskeletal Diseases | – | – | – | – | 1.054* [1.002, 1.109] | 0.986 [0.947, 1.027] |
| Malformations/Abnormalities | – | – | – | – | 1.185* [1.019, 1.378] | 0.890 [0.778, 1.018] |

*Note:* Values are hazard ratios with 95% confidence intervals in brackets. Early PE = Early-onset pre-eclampsia (<34 weeks); Late PE = Late-onset pre-eclampsia (≥34 weeks). '—' indicates the variable was not included in that model. * = p<0.05.
